# Trends in Developmental Milestone Attainment Among Israeli Children, 2019-2024

**DOI:** 10.1101/2025.09.30.25336963

**Authors:** Irena Girshovitz, Guy Amit, Inbal Goldshtein, Pinchas Akiva, Dalit Landesman Milo, Deena R. Zimmerman, Ravit Baruch, Meital Avgil Tsadok, Yair Sadaka

## Abstract

**Background:** Monitoring developmental milestones is essential for identifying at-risk populations and guiding effective intervention programs. Our previous report analyzed milestone attainment among Israeli children from 2016 to 2020. Using a similar methodology, this study examines trends in developmental milestone attainment within Israeli population from 2019 to 2024.

**Methods:** The dataset included over 1 million children with more than 5 million developmental assessments conducted at Maternal Child Health Clinics (MCHCs) across Israel. These assessments were used to calculate failure rates across four developmental domains: language, social, gross motor, and fine motor skills, along with parental concern about child development. Data were stratified by maternal characteristics, such as age, education, employment, and marital status, and by family sociodemographic factors, including ethnicity, degree of Haredi observance and socioeconomic status. We applied multivariable logistic regression to analyze the impact of different sociodemographic factors on the odds of failure to attain milestones, while controlling for confounding.

**Results:** A mild overall increase in milestone failure rates was observed from 2019 to 2023. Disparities based on maternal characteristics, particularly lower education, divorce status, unemployment, advanced maternal age, and immigration background, widened notably during periods of national disruption (COVID-19 and the war). In contrast, gaps associated with ethnicity and socioeconomic status remained stable or narrowed. Parental concern increased across most groups, particularly among Bedouin parents, suggesting growing awareness.

**Conclusions:** Maternal-level characteristics were strongly associated with elevated developmental risk observed from 2019 to 2023, and may be more sensitive to national crises. The findings underscore the importance of strengthening parental awareness, prioritizing early identification of high-risk families, and maintaining access to developmental services during emergencies.

## Background

### Introduction

Early childhood is a critical period for child development, as delays in milestone attainment may indicate underlying medical or developmental conditions. Developmental surveillance has become a global standard, supported by initiatives like the American Academy of Pediatrics’ “Learn the Signs. Act Early” program [1]. In Israel, Maternal Child Health Clinics (MCHCs), known as “Tipat Halav”, provide routine developmental monitoring through scheduled visits from birth to six years of age [2].

A previous national report (2016-2020) [3] identified concerning trends, including increasing rates of failures in milestone attainment across all developmental domains, with the most notable rise observed in the language domain. It also emphasized the elevated risks faced by children from socioeconomically disadvantaged backgrounds, particularly those with unemployed mothers or from minority ethnic groups. These findings underscored the urgent need for targeted interventions and systemic support to reduce disparities and promote optimal developmental outcomes.

This updated report (2019-2024) follows a similar methodology and incorporates data collected during periods of significant disruption, including the COVID-19 pandemic, and the war that began in October 2023. In February 2023, a policy change modified the timing of assessment for certain developmental milestones and introduced several new ones. This change affected the evaluation schedule, potentially influencing failure rates and complicating comparisons with previous years.

Despite these challenges, this report aims to provide a comprehensive analysis of recent developmental trends, identify areas requiring further intervention, and inform policy decisions that support equitable child development across Israel.

## Methods

### Study Population

The study population included 1,158,536 children who attended 494 MCHCs across Israel between October 2018 and September 2024. More than 5 million developmental evaluations were conducted, covering four domains: language, social, gross motor, and fine motor skills. All evaluations followed consistent age thresholds for assessing the severity of failures in milestone attainment [4]. These thresholds remained unchanged during the study period, ensuring comparability over time.

Three major disruptions affected the data collection process:

1. **Covid-19 pandemic:** Due to changes in the surveillance policy and service availability between March and October 2020, surveillance visits to MCHCs were irregular, primarily as a result of constraints imposed by the COVID-19 pandemic. These disruptions may have contributed to specific changes observed during this period.
2. **Policy update**: In early 2023, a Ministry of Health directive introduced new developmental milestones and modified the timing of assessment for some of the existing ones [5]. To ensure consistency, the primary analysis in this report included only milestones whose evaluation timing remained unchanged from previous guidelines. In addition, we present the results in two separate time periods: from the beginning of October 2018 to the end of February 2023, and from the beginning of March 2023 to the end of September 2024. This division was necessary due to the change in the assessment method.
3. **Impact of conflict**: The war that started in October 2023 caused substantial disruptions to routine evaluations during its initial months. To reduce the effect of these irregularities, data for each reporting year were aggregated from October through September of the following year. Accordingly, 2024, the final year of the analysis, spans from October 2023 to September 2024 and includes evaluations conducted during the conflict.

### Measured Developmental Outcomes

This report employed the Tipat Halav Israel Surveillance (THIS) developmental scale [4], which defines milestone attainment age thresholds at the 75th, 90th, and 95th percentiles of the population. Following the methodology of the previous report, “failure” was defined as the inability to attain a milestone by the age at which it is attained by 95% of the children. For each developmental domain, language, social, gross motor, and fine motor, we calculated the annual percentage of children failing to attain at least one milestone, considering only the first failure per domain per year. Self-reported parental concern regarding their child’s development was also recorded and analyzed.

### Statistical analysis

We report the annual rate of children who did not attain developmental milestones, both overall and stratified by sociodemographic factors, along with 95% confidence intervals for proportions. Data were presented at the national level and by geographic subdivisions. Missing values in categorical sociodemographic variables were treated as a separate category.

Key sociodemographic variables included maternal level of education, ethnic group, country of birth, employment status, marital status, and age at delivery. We also included an analysis of the association between ultra-Orthodox (Haredi) population concentration in the residential area and milestone attainment rates. The child’s sex and age at assessment were also included in the analyses.

Socioeconomic clusters and Haredi Level were extracted from the Points measures [6]. Haredi Level reflects the proportion of the Haredi population in a given area and is categorized as: none, low, medium, high, and very high. For our analyses, we defined ‘none’ as representing non-Haredi areas, high’ and ‘very high’ as Haredi areas, and ‘low’ and ‘medium’ as heterogeneous areas. Descriptive statistics for baseline population characteristics were expressed as percentages. Comparisons between sociodemographic subgroups were conducted using χ2 tests, with Bonferroni correction applied for multiple comparisons. Trends in milestone failure rates over time were assessed using the Cochran–Armitage trend test.

Multivariable logistic regression was used to estimate the association between each explanatory variable and the odds of failing to attain developmental milestones, adjusting for potential confounders. This analysis was repeated separately for each developmental domain. Variance inflation factor (VIF) was used to assess multicollinearity among explanatory variables (results are presented in Supplementary Table S1). The analyses were performed using Python programming language version 3.9 (Python Software Foundation) and R version 4.4.2 (R Project for Statistical Computing).

## Results

The main characteristics of the cohort are presented in Table 1. We compared the number of children aged 0-5 years in the study cohort with population estimates from the Israeli Central Bureau of Statistics (CBS). The cohort includes approximately 73% of all Israeli children during the study period and is fairly representative of the national population. However, there is an overrepresentation of minority groups: 76% of the Israeli-Arab children and 93% of the Druze children are included, compared to 59% of the Israeli-Jewish children. The full results of this comparison can be found in Supplementary Table S2.

**Table 1.**
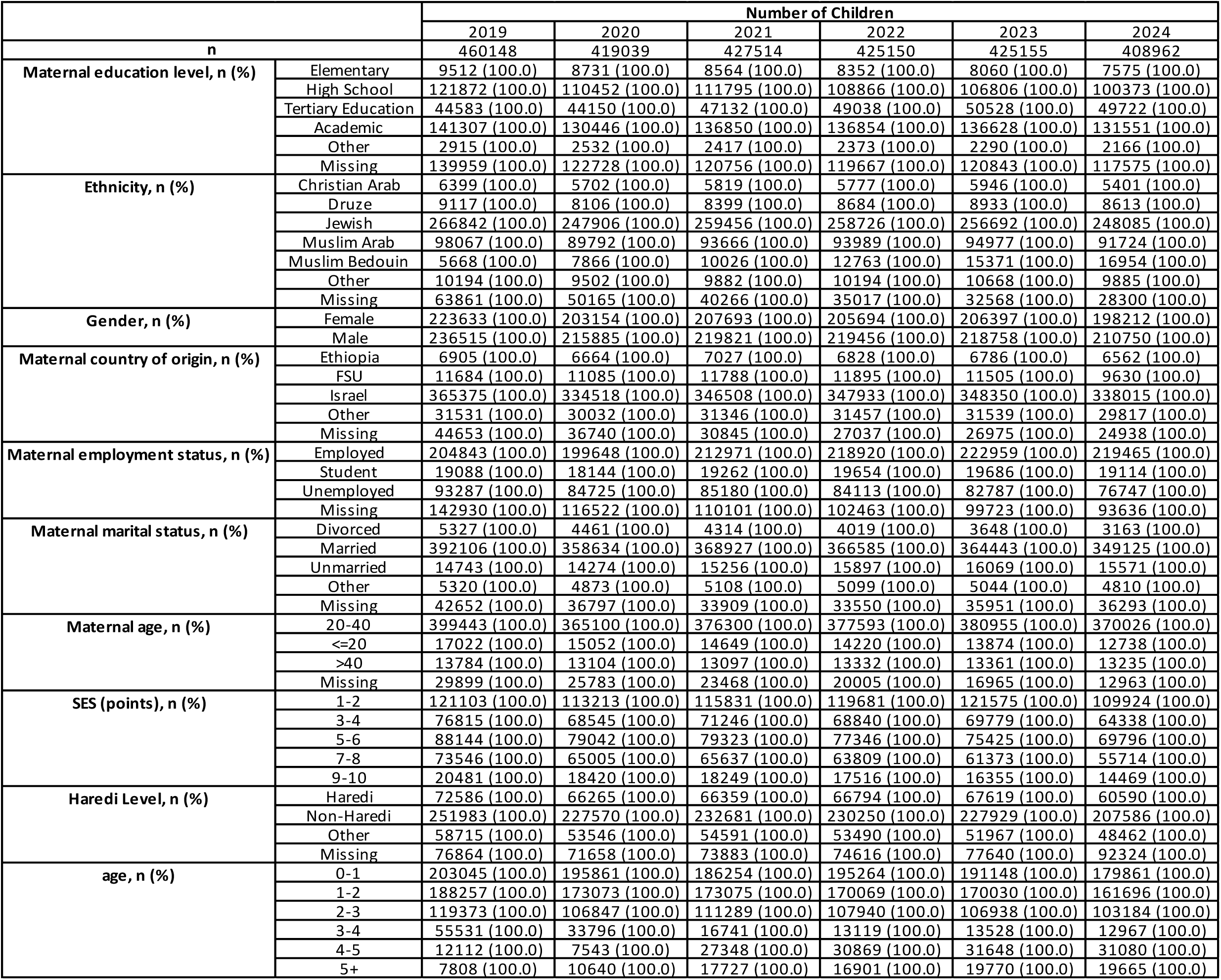
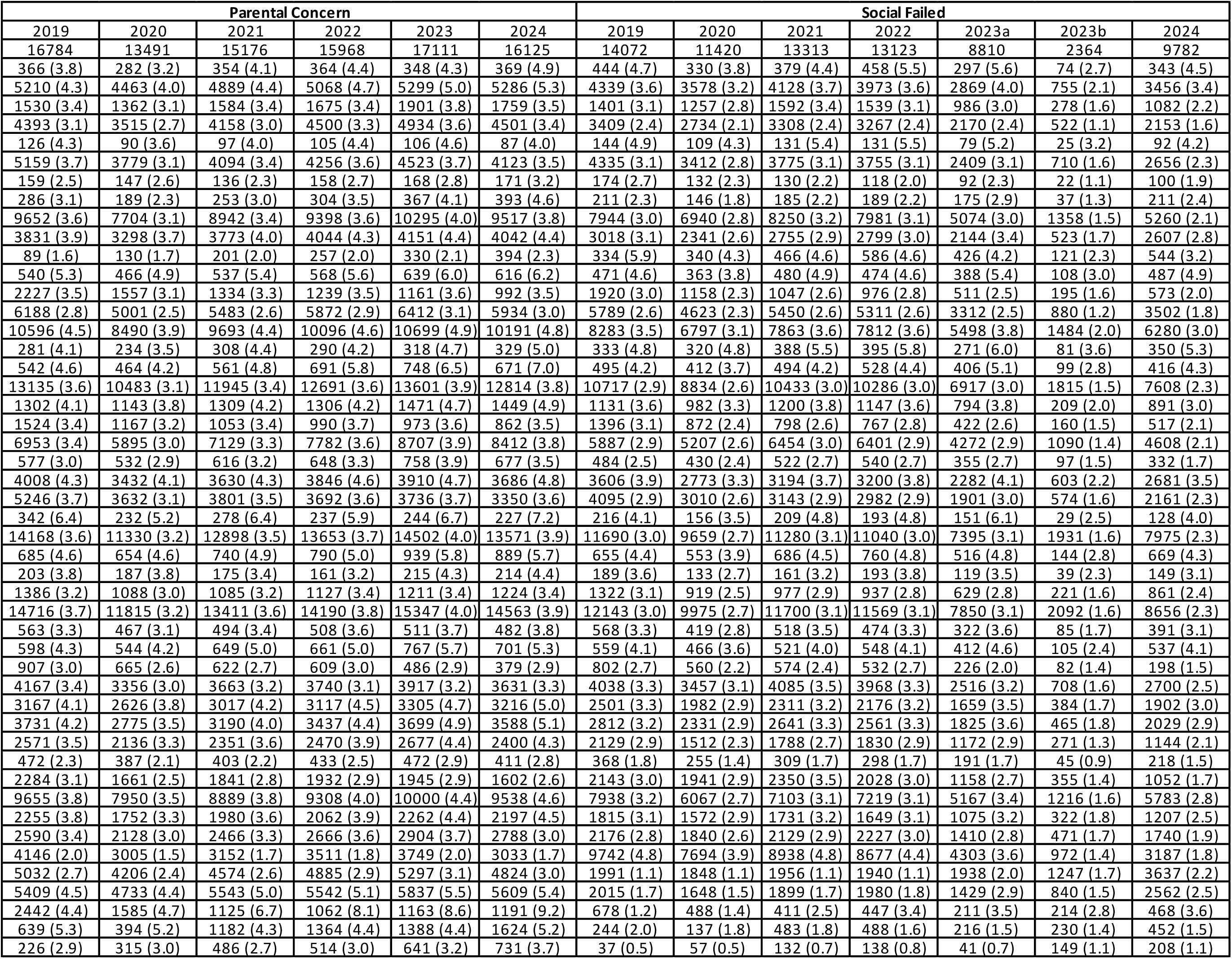
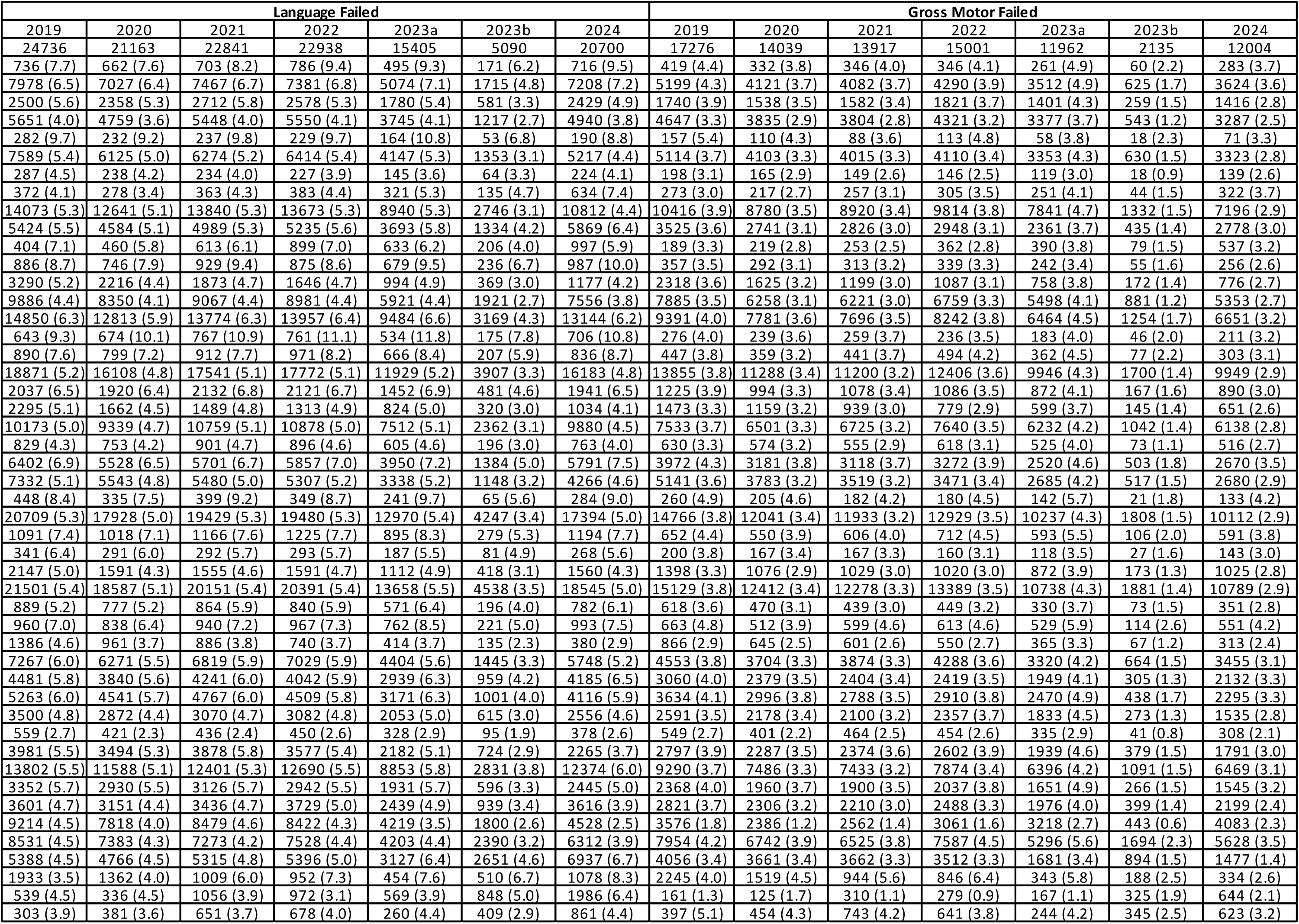
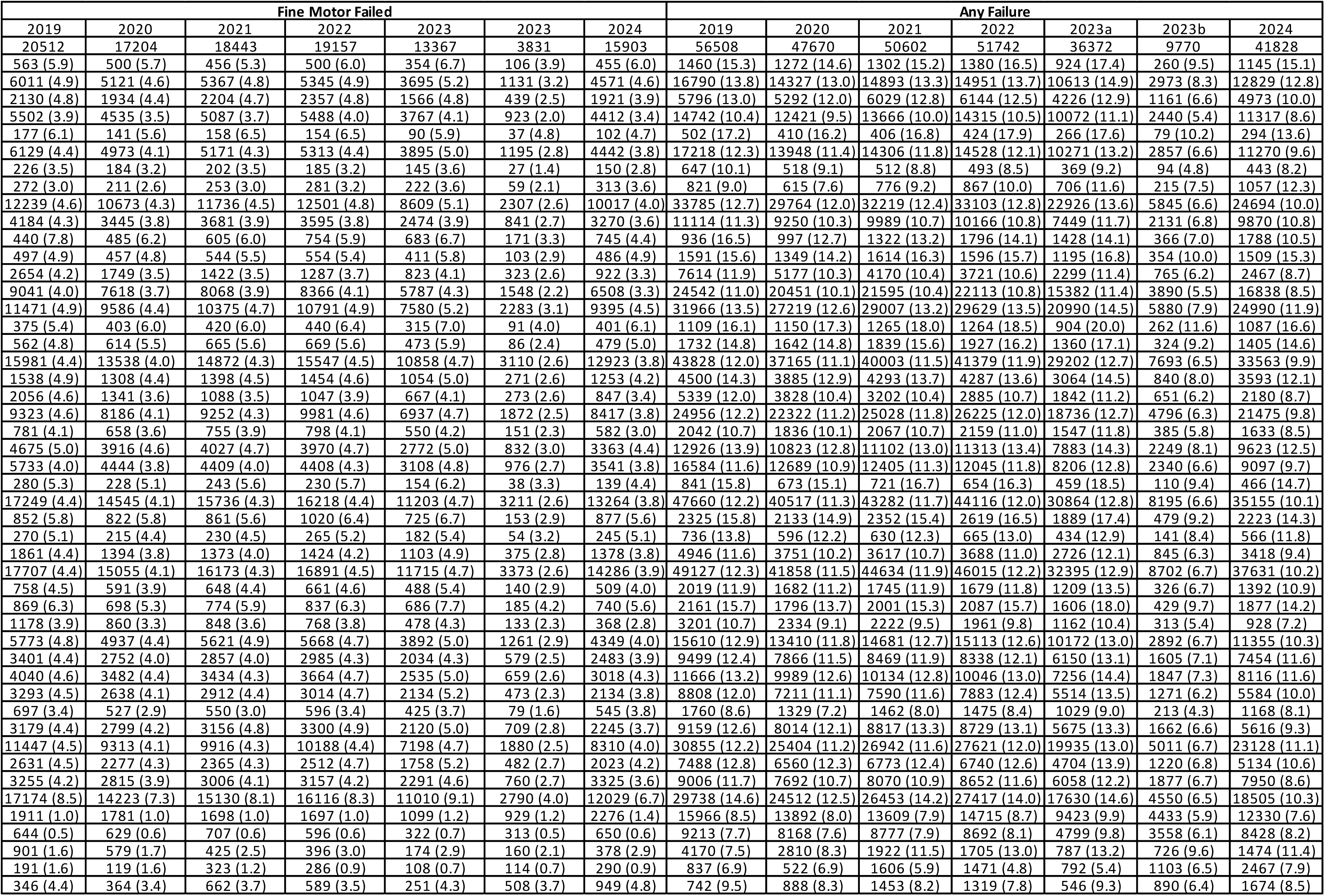
Characteristics of study population by year, stratified by parental concern and failure in language, social, fine motor, gross motor, or any developmental domain. In the religion variable, mothers whose origin was reported “Circassian”, “Other Muslim”, “Other Christian” or “Other” were all sub-grouped as “Other”. In the maternal education level “Other” includes “No education” and “Studies in Yeshiva”. In the birth country variable, “Other” includes children whose mothers were reported as born in Europe, Other, North America, Africa, Asia, South and Central America, Australia, and Oceania. In marital status, ‘widower’ and ‘other’ were reported as ‘Other’.

## General trends

In contrast to the previous report, which showed a significant increase over the reported years, the current analysis (2019–2023) reveals only a mild upward trend in the rate of failure to attain developmental milestones across all four domains, as illustrated in Figure 1.

**Figure 1.**
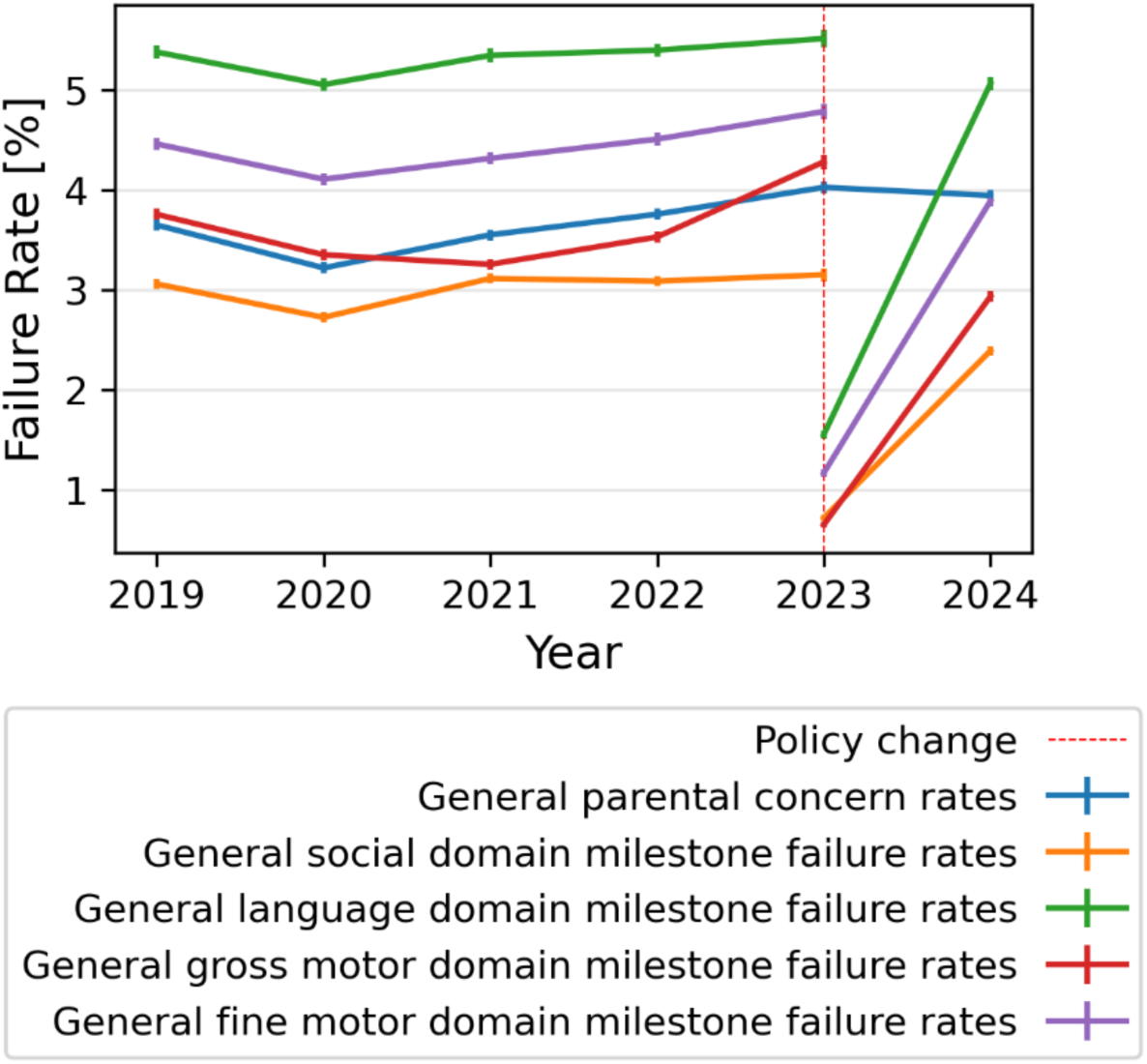
Temporal trends in child development domains and in parental and nurse’s report of concern for child development.

The milestone attainment failure rate in the gross motor domain ranged from 3.75% in 2019 (95% CI: [3.70, 3.81]), to 4.27% in 2023 [4.20, 4.35]. In the fine motor domain, the failure rate was 4.46% in 2019 [4.40, 4.52], and 4.78% in 2023 [4.70, 4.86]. The social domain showed a failure rate of 3.06% in 2019 [3.01, 3.11] and 3.15% in 2023 [3.09, 3.22]. As in the previous report, the highest failure rates were observed in language milestones: 5.4% in 2019 [5.31, 5.44] and 5.51% in 2023 [5.43, 5.60].

In 2023 (after policy change), failure rates across all developmental domains appeared lower compared to the period before the policy change, but began to rise again in 2024.

Parental concern also increased over time, from 3.65% in 2019 [3.59, 3.70] to 3.94% in 2024 [3.88, 4.00], in contrast to the decrease reported in the previous analysis.

## Maternal characteristics and Sociodemographic correlates of child development

### Maternal education

Figure 2 presents the temporal trends in developmental outcomes stratified by maternal education level. Consistent with our previous findings, failure rates across all developmental domains decrease as the maternal education level increases. This trend is most pronounced in the language and social domain (Figures 2b, 2c), and less evident in the gross motor domain (Figure 2d).

**Figure 2.**
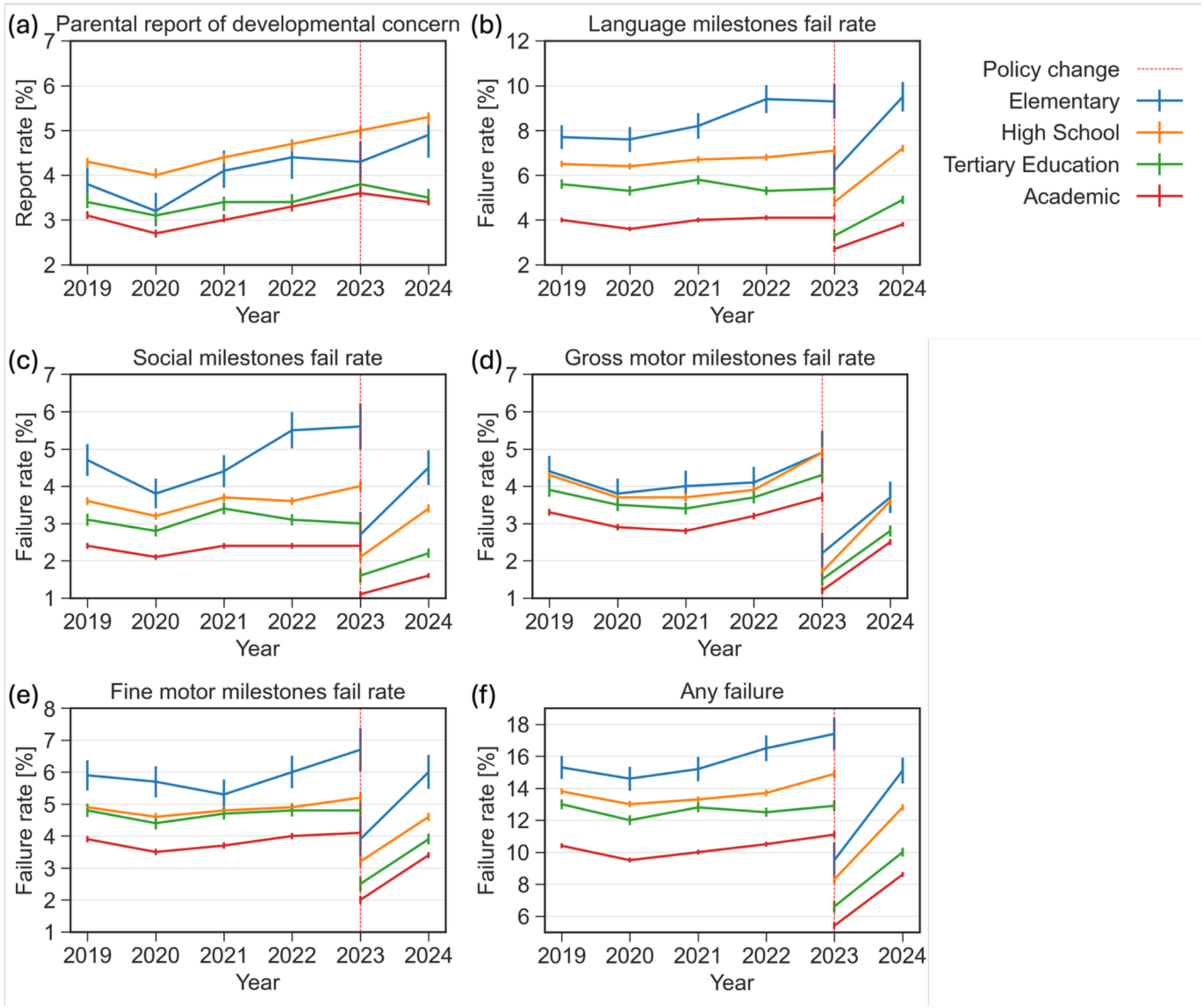
Reports of concern and failure rates in various developmental domains stratified by maternal education level between 2019-2024. (a) Parental report of developmental concern (b) Failure rate in the language domain (c) Failure rate in the social domain (d) Failure rate in the gross motor domain (e) Failure rate in the fine motor domain (f) Failure rate in any developmental domain

In the language, social and fine motor domains, failure rates have been increasing more rapidly among children whose mothers have lower levels of education, thus widening the gap between educational groups (Figures 2b, 2c, 2e). For example, the percentage of children failing to attain language milestones whose mothers had only elementary education rose from 7.7% in 2019 to 9.3% in 2023, prior to the policy change. After the change, the failure rate was 6.2% and increased to 9.5% in 2024. In contrast, among children whose mothers had an academic education, the failure rate remained relatively stable, at 4% in 2019 and 4.1% in 2023, prior to the policy change (p<0.001). After the policy change, the rate was 2.7%, followed by a slight rise to 3.8% in 2024.

In the fine motor and social domains, failure rates in 2019 were 5.9% and 4.7%, respectively, among children whose mothers had only elementary education, compared to 3.9% and 2.4% among children whose mothers had academic education (p < 0.001). By 2023 (prior to policy change), these rates rose to 6.7% (fine motor) and 5.6% (social) in the elementary education group, and to 4.1% and 2.4%, respectively, in the academic education group (p < 0.001).

In 2023 (post policy change), the fine motor failure rate for children of academically educated mothers was 2.0%, while for children of mothers with only elementary education, the rate was 3.9%. In 2024 the rates were 3.4% and 6%, respectively (p<0.001). In the social domain, the failure rate among children of academically educated mothers was 1.1% in 2023 and 1.6% in 2024, while among children whose mothers had elementary education, the rate was 2.7% in 2023 and 4.5% in 2024 (p<0.001).

### Ethnic group

Figure 3 illustrates trends in developmental outcomes across ethnic and religious groups, including Jews, Muslim Arabs, Christian Arabs, Druze, and Bedouin. Among these groups, Bedouin children exhibited higher failure rates in several domains: language (6.2% in 2023 (pre-policy change), p<0.001), social (4.2% in 2023 (pre-policy change), p<0.001; 3.2% in 2024, p<0.001), and fine motor (6.7% in 2023 (pre-policy change), p<0.001; 4.4% in 2024, p<0.001) milestones (Figure 3b, 3c, 3e). Despite these higher failure rates, parental concern within the Bedouin community was the lowest among all groups (Figure 3a), suggesting lower awareness of potential developmental delays. However, over the past six years, this population (as well as the Druze population) has shown the most significant increase in reports of parental concern.

**Figure 3.**
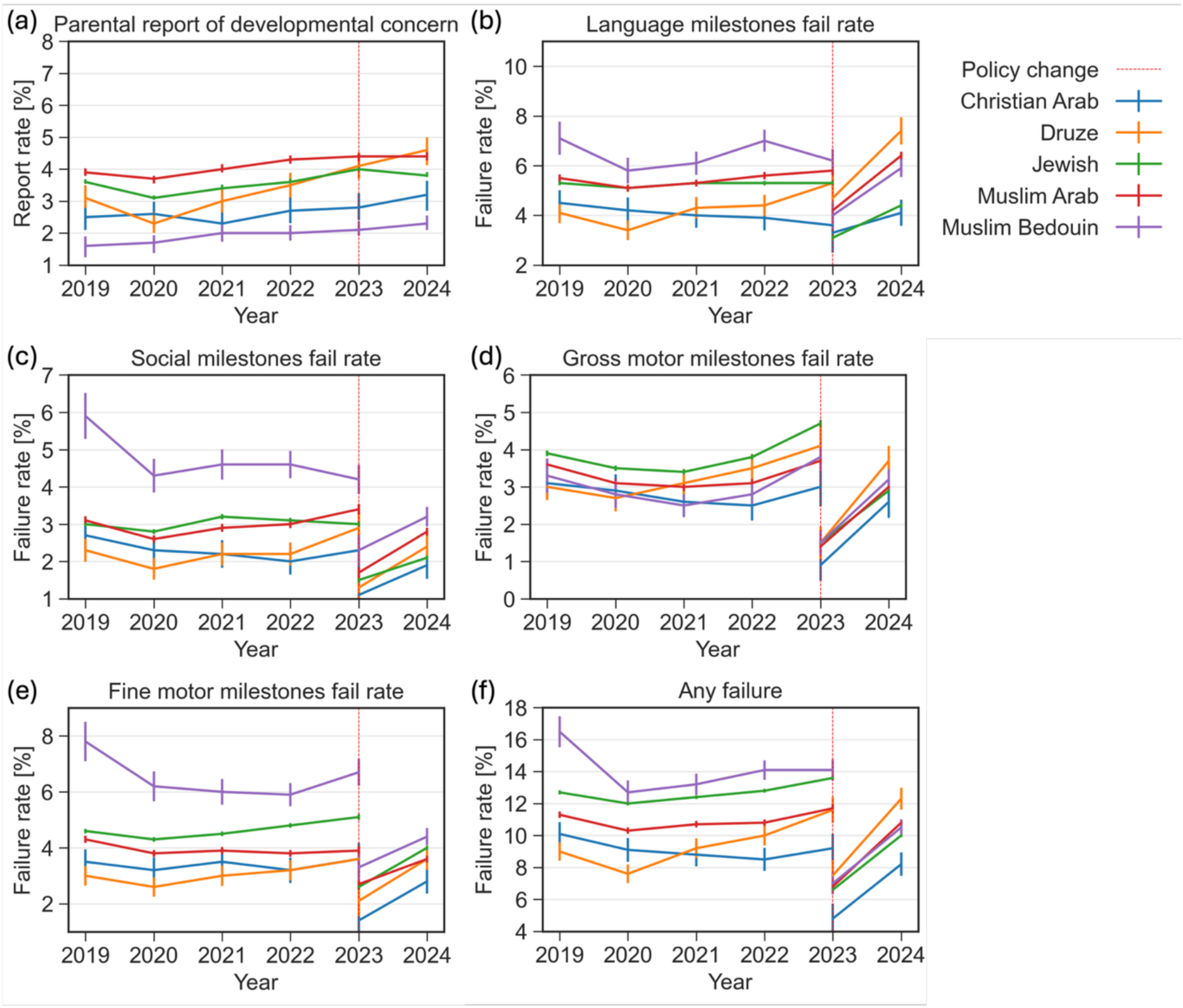
Reports of concern and failure rates in various developmental domains stratified by ethnic and religious groups between 2019-2024. (a) Parental report of developmental concern (b) Failure rate in the language domain (c) Failure rate in the social domain (d) Failure rate in the gross motor domain (e) Failure rate in the fine motor domain (f) Failure rate in any developmental domain

### Maternal birth country

Figure 4 presents trends in developmental outcomes stratified by maternal birth country. Failure rates in social, language, and fine motor milestones were higher among children whose mothers immigrated to Israel from Ethiopia and, to a lesser extent, from the Former Soviet Union (FSU).

**Figure 4.**
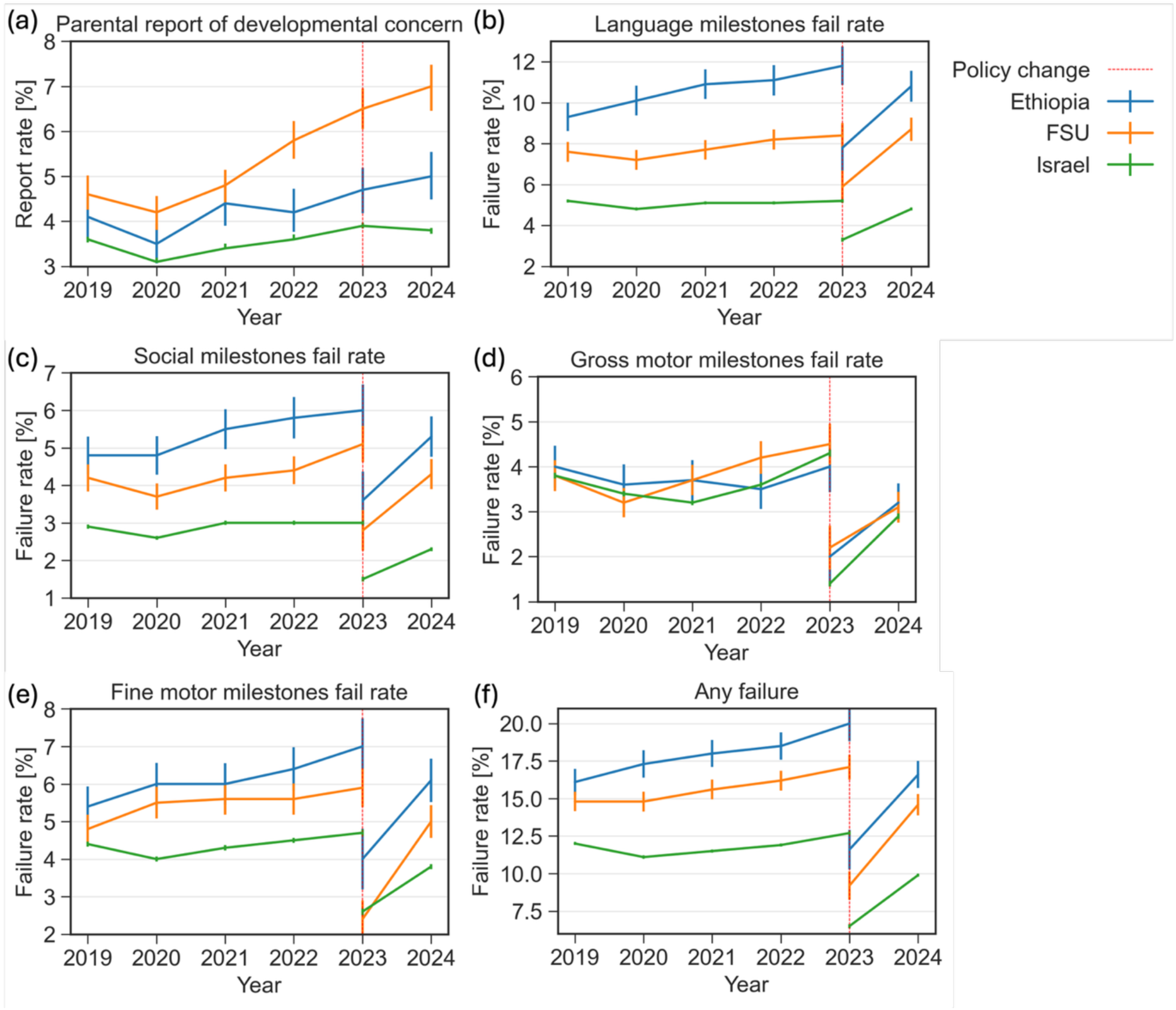
Reports of concern and failure rates in various developmental domains stratified by maternal birth country between 2019-2024. (a) Parental report of developmental concern (b) Failure rate in the language domain (c) Failure rate in the social domain (d) Failure rate in the gross motor domain (e) Failure rate in the fine motor domain (f) Failure rate in any developmental domain

Parental concern regarding child development increased between 2019 and 2023 (prior to policy change) among mothers born in Israel (P<0.01) and those from the FSU (p<0.001), with greater increase observed among immigrant mothers. The rate of parental concern was highest among mothers who were born in the FSU (7% in 2024) compared to mothers born in Israel or other countries (3.8%-5%; p<0.001).

### Maternal employment status

Supplementary Figure 1 illustrates the association between maternal employment status and developmental outcomes. Across all four domains, milestone failure rates were higher among children of unemployed mothers, compared with children of employed or student mothers.

This difference was most pronounced in the language domain, where failure rates among children of unemployed mothers were 7.2% in 2023 prior to policy change and 7.5% in 2024 (P<0.001), compared to 5.1% in 2023 and 4.5% in 2024 among children of employed mothers (p<0.001). Similarly, parental concern regarding developmental delays was reported more frequently by unemployed mothers than by employed mothers or students.

### Maternal marital status

Supplementary Figure 2 presents the association between maternal marital status and developmental outcomes. In 2024, married mothers were less likely to express concerns regarding their child’s development (3.9%, p<0.001), compared to unmarried (5.7%, p<0.001) or divorced (7.2%, p<0.001) women (Supplementary Figure 2a). Similar patterns were observed in objective failure to attain developmental milestones across all four domains, according to the THIS scale (Supplementary Figure 2b-2e).

### Socioeconomic Status (SES)

Figure 5 illustrates developmental milestone attainment across various SES cluster pairs. Until 2023, prior to policy change, clusters 9–10 consistently exhibited the lowest failure rates in all developmental domains, according to the Points measures.

**Figure 5.**
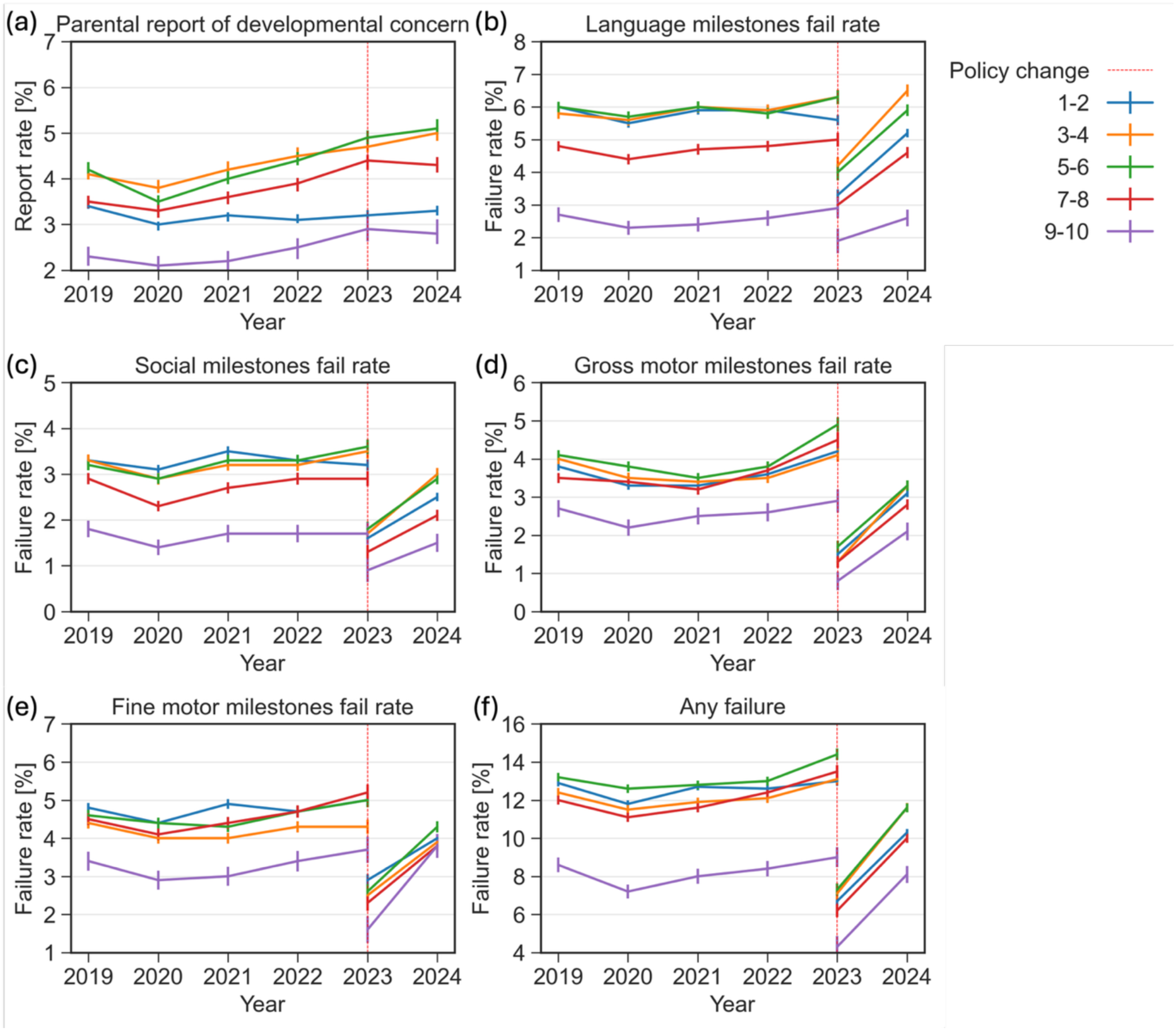
Reports of concern and failure rates in various developmental domains stratified by CBS-based socioeconomic status between 2019-2024. (a) Parental report of developmental concern (b) Failure rate in the language domain (c) Failure rate in the social domain (d) Failure rate in the gross motor domain (e) Failure rate in the fine motor domain (f) Failure rate in any developmental domain

In 2024, however, the fine motor failure rate in clusters 9-10 was 3.7%, narrowing the gap with clusters 1–8, which ranged from 3.6% to 4.4%.

Parents in clusters 9–10 reported lower concern regarding their child’s development (p<0.001).

### Haredi Level

Figure 6 illustrates the association between Haredi level and developmental milestone attainment. Parents living in non-Haredi areas expressed higher levels of concern regarding their child’s development compared to those in Haredi areas (4.6% vs. 2.6% in 2024, respectfully; p<0.001).

**Figure 6.**
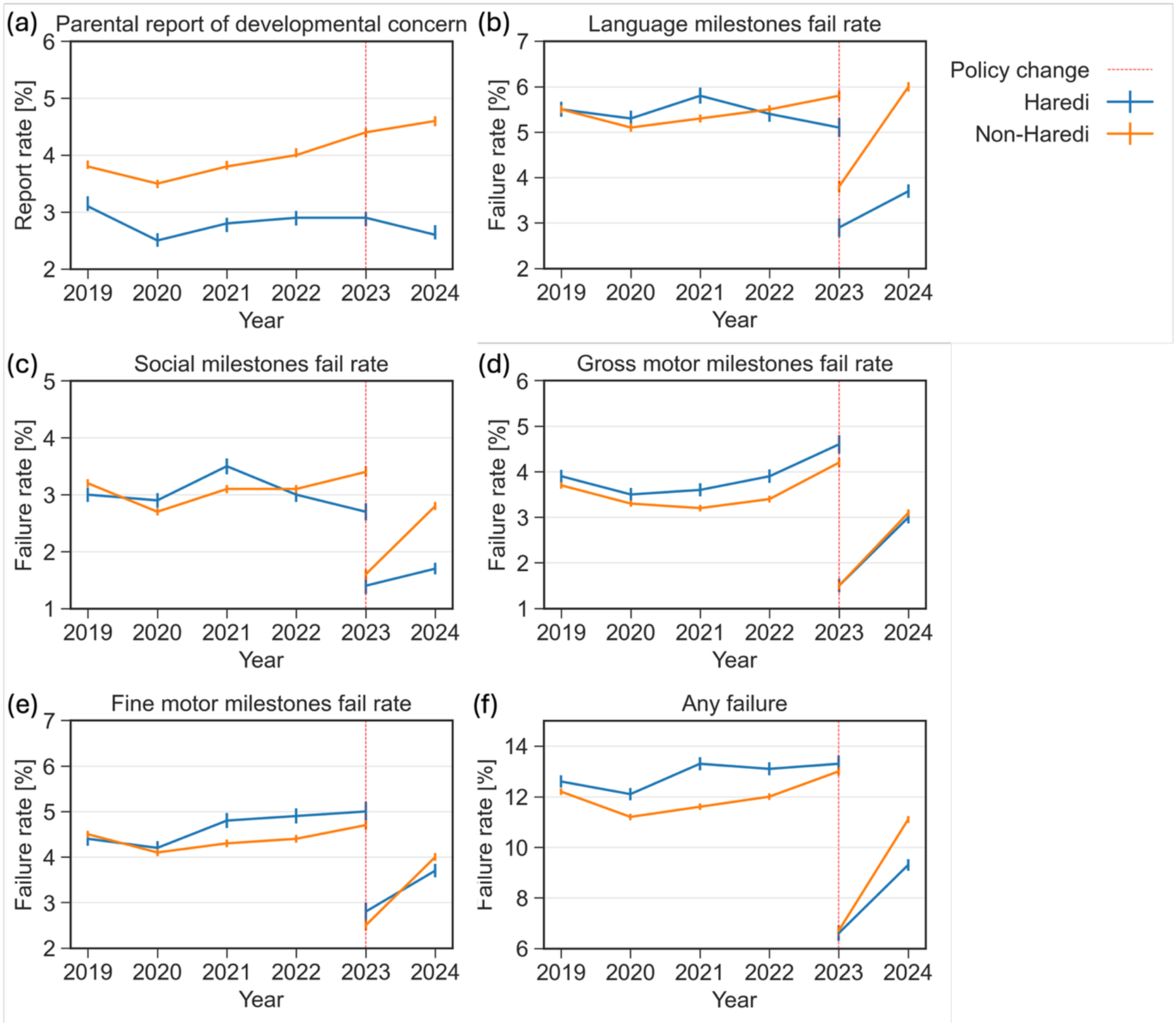
Reports of concern and failure rates in various developmental domains stratified by Haredi level between 2019-2024. (a) Parental report of developmental concern (b) Failure rate in the language domain (c) Failure rate in the social domain (d) Failure rate in the gross motor domain (e) Failure rate in the fine motor domain (f) Failure rate in any developmental domain

**Figure 6.**
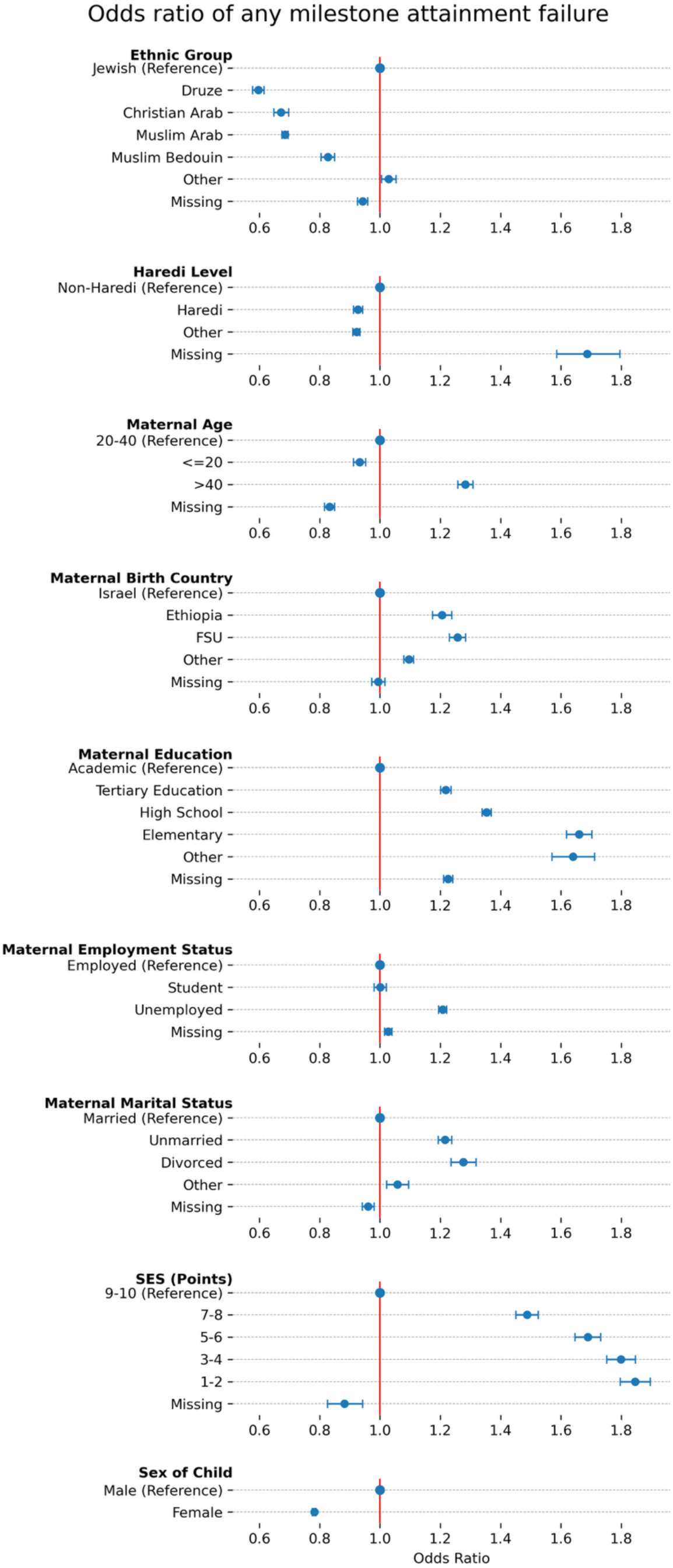
Multivariable analysis for any failure for 2019-2022, where x axis is the odds ratio and y axis is the demographic variables.

In the social domain, failure rates remained stable from 2019 to 2023 (prior to policy change), with a slight reduction among Haredi children and a slight increase among non-Haredi children (p<0.001). Conversely, in the language domain, Haredi children experienced a decrease in failure rates from 5.1% in 2023, prior to policy change to 3.7% in 2024, while children in non-Haredi areas showed an increase from 5.8% to 6% (p<0.001).

In contrast, for the gross and fine motor domains, Haredi children consistently exhibited higher failure rates between 2020 and 2023 (prior to policy change).

By 2024, the gaps in the language and the social domains had widened, as the Haredi population exhibited a less pronounced increase in failure percentages.

### Sex of child

As in the previous report, failure rates were higher among males, compared to females, across all developmental domains (Supplementary Figure 3), although the difference was not statistically significant in the gross motor domain. The largest disparity between sex groups was observed in the language domain. For example, in 2024, 6.2% of assessed males failed to attain language milestones, compared to 3.8% of assessed females, P<0.001).

### Maternal age

Supplementary Figure 4 presents data on parental concern and failure rates across developmental domains, stratified by maternal age. Consistent with previous findings, women who gave birth at age 40 or older reported higher levels of concern regarding their child’s development compared to those who gave birth between ages 20-40 or before age 20 (p<0.01 for 2019–2023). In 2024, the difference was even more pronounced when comparing mothers over 40 to those aged 20–40 (p<0.001). Additionally, children of adolescent mothers (<20) showed increased rates of unmet developmental milestones, especially in social and language domains, between 2023 and 2024.

Similar patterns were observed in milestone attainment failure rates across all four domains (Supplementary Figure 4b-4e).

Additional sub-analysis by geographical districts is available online (in Hebrew) [7].

## Association between sociodemographic variables and developmental milestone attainment

Mutually adjusted odds ratios (ORs) for failure to attain any milestone, as well as for domain-specific failures in 2019-2023 prior to policy change are presented in Figure 6 and Supplementary Figures 5-8, respectively. Corresponding analyses for 2023-2024 (after policy change) are shown in Figure 7, and Supplementary Figures 9-12, respectively.

**Figure 7.**
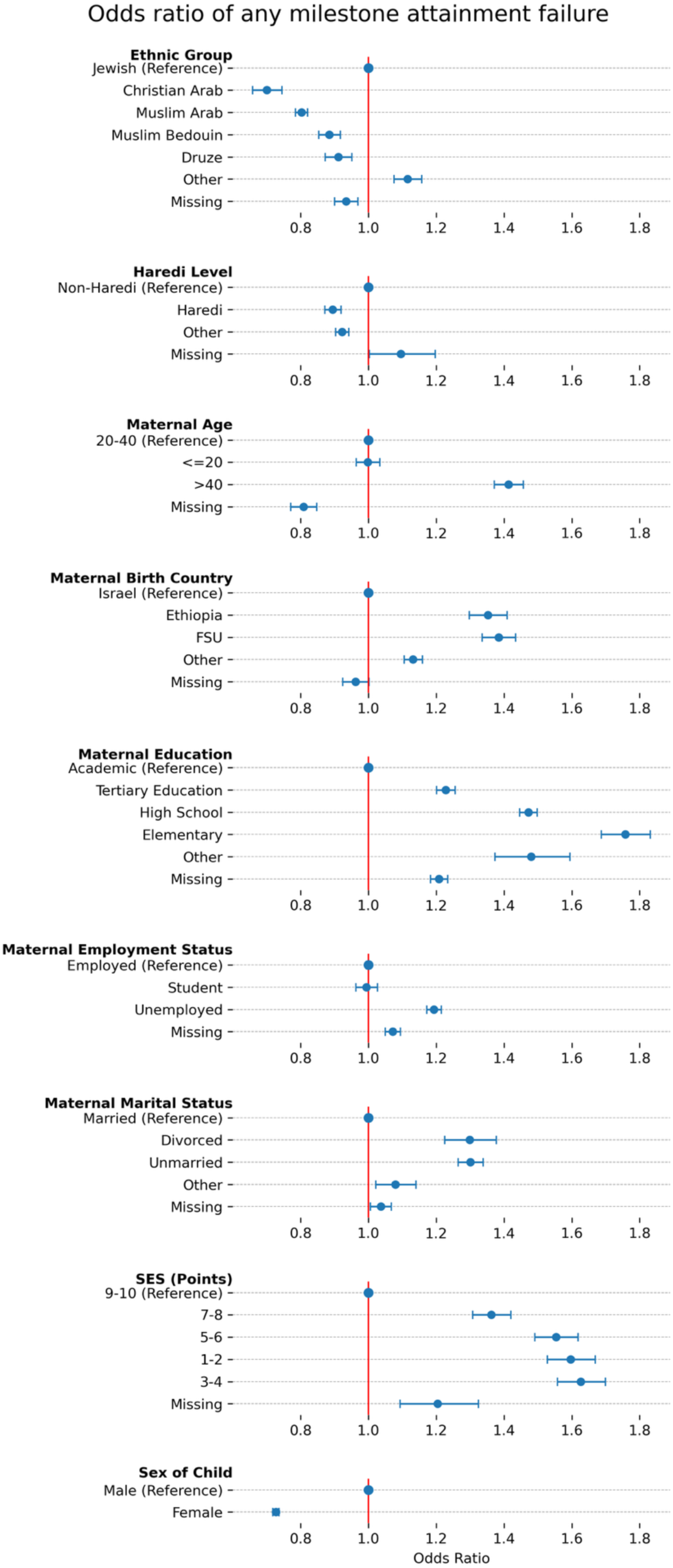
Multivariable analysis for any failure for 2023-2024, where x axis is the odds ratio and y axis is the demographic variables.

During 2019-2023, higher odds of failure to attain any developmental milestone was observed among children whose mothers were divorced (OR=1.28 [1.23, 1.32], P<0.001) or unmarried (1.22 [1.19, 1.24], P<0.001), unemployed (1.21 [1.19, 1.22], P<0.001), immigrant from the FSU (1.26 [1.23, 1.28], P<0.001) or Ethiopia (1.21 [1.17, 1.24], P<0.001), have elementary (1.66 [1.62, 1.70], P<0.001) or high school (1.35 [1.34, 1.37], P<0.001) education, or were aged over 40 years at the time of childbirth (1.28 [1.26, 1.31], P<0.001).

Children from lower socioeconomic status (SES) clusters also had elevated risk, with odds ratios ranging from 1.49 to 1.85. The increased risk was mostly evident in the language (ORs ranging from 1.43 to 2.45), social (1.64-2.23) and fine motor (1.43-1.73) domains.

In 2023-2024, higher risk was observed in children to mothers who are divorced (1.3 [1.23, 1.38], P<0.001) or unmarried (1.30 [1.26, 1.34], P<0.001), unemployed (1.19 [1.17, 1.22], P<0.001), immigrant from the FSU (1.38 [1.34, 1.43], P<0.001) or Ethiopia (1.35 [1.30, 1.41], P<0.001), have elementary (1.76 [1.69, 1.83], P<0.001), high school (1.47 [1.45, 1.50], P<0.001) or tertiary (1.23 [1.20, 1.26]) education, or were aged over 40 years at the time of childbirth (1.41 [1.37, 1.46], P<0.001).

Children from lower SES clusters also had elevated risk, with odds ratios ranging from 1.36 to 1.62. As in the earlier period, the language, social and fine motor domains showed the most pronounced disparities.

## Discussion

This report offers a unique longitudinal perspective on child development indicators in Israel, utilizing data from the MCHC system from 2019 to 2024. The 2023 policy update, which revised milestone assessment timing and introduced new developmental measures, required separate analyses for the periods 2019–2023 and 2023–2024. The findings support evidence-based policymaking and provide valuable insights into the relationship between sociodemographic factors and child development.

### General developmental trends

In contrast to the previous report, which showed a significant increase over the reported years, the current analysis (2019-2023) reveals only a mild upward trend in the rate of failure to attain developmental milestones across all four domains. Parental concerns regarding child development increased steadily from 2019 to 2024, reversing the downward trend observed between 2016 and 2020 [3]. Notably, Bedouin parents remained the least likely to report concern overall, yet both Bedouin and Druze parents showed the largest increases in concern over time. This trend may reflect growing awareness of developmental challenges, which could positively influence early intervention efforts.

Our comparison of developmental milestone attainment between boys and girls suggests that using a single evaluation scale may be insufficient, as their developmental trajectories likely differ. This underscores the potential need for sex-specific developmental assessment scales [8].

These observed trends across different populations may, in part, be related to recent changes in staff training, which place greater emphasis on language-related aspects and parental guidance during developmental evaluations.

### Maternal characteristics

Maternal characteristics are key factors in children’s development outcomes, particularly in the attainment of language and social milestones. Throughout the reporting period, notable differences were observed between children whose mothers had favorable conditions, such as higher education, steady employment, and supportive family structures, and those whose mothers faced more challenging circumstances. These disparities were most pronounced in language and social development, and they became more pronounced between 2019 and 2023 [9,10].

In 2024, following both changes in screening protocols and the broader impact of wartime disruptions, these developmental gaps further expanded in most domains. The disparities associated with maternal education, marital status, age, and country of origin all showed expanded developmental gaps, particularly in language and social development. Children of mothers with lower education experienced the sharpest growth in failure rates, as did children of divorced mothers and those born to older mothers (aged 40 and above). Similarly, children of Ethiopian-born mothers continued to exhibit the highest failure rates, with no significant narrowing of the ethnic-origin gap. The only exception to this overall trend was observed in maternal employment: although children of unemployed mothers generally had higher failure rates, no meaningful increase in the disparity between employed and unemployed groups was detected in 2023-2024, suggesting a relative stabilization in that dimension.

### Socioeconomic and Ethnic Backgrounds

In contrast to the widening gaps associated with maternal characteristics, disparities related to socioeconomic and ethnic background showed a relatively stable or narrowing trend throughout the reporting years. SES remained a significant factor, with the lowest failure rates consistently observed among children from the highest SES clusters (9-10), a pattern widely supported in the literature [11,12]. However, these gaps did not expand over time. Notably, in 2023-2024, a rise in failure rates was recorded among children from SES clusters 9-10, particularly in fine and gross motor domains, contributing to a partial reduction in disparities between SES groups.

Ethnic differences also revealed a gradual narrowing over the years. While the Bedouin population continued to show the highest failure rates in 2019-2023, especially in language and social domains, followed by the Muslim Arab and Jewish populations, the overall trend pointed toward convergence. The Christian Arab and Druze populations maintained the lowest failure rates.

Over the reporting period, a gradual widening of gaps was observed in language and social development between children from Haredi and non-Haredi communities, with this trend further intensifying in 2024. Parents residing in non-Haredi areas consistently reported higher levels of developmental concern compared to those in predominantly Haredi regions. Consistently, although the differences in failure rates between the two groups were relatively modest in the language and social domains, a slight increase in failure rates was noted among children from non-Haredi populations compared to their Haredi peers. In contrast, in both fine and gross motor development, children from Haredi backgrounds exhibited consistently higher failure rates throughout most years of the report. In 2023-2024, the increase was more pronounced among non-Haredi children, particularly in language, social, and fine motor domains.

### Strengths and Limitations

This study benefits from the unique infrastructure of the Tipat Halav system, enabling large-scale, longitudinal tracking of child development across Israel. By focusing on milestones with unchanged assessment timing, the methodological approach ensures consistency and comparability over time, including in the context of the 2023 protocol update. To account for changes introduced in that update, results for 2023–2024 are reported separately.

However, several limitations must be acknowledged. Approximately 30% of children attending non-MOH MCHCs were not included. Nevertheless, both prior research [4] and our own validation suggest this population is broadly representative of the national demographic. Additionally, geographic and cultural variability in assessment practices may have introduced inconsistencies in data collection. Furthermore, the variables obtained from Points, such as SES and Haredi Level, are aggregated at the residential area level, rather than at the individual level.

Irregular MCHC visits during the COVID-19 pandemic alongside an increased reliance on parent-reported assessments, and the effect of resident evacuations, postponed MCHC visits, and parental reports during the 2023 war, may have affected the accuracy and completeness of developmental milestone tracking. Furthermore, the 2023 protocol update may have introduced unintended bias by increasing emphasis on developmental assessments, even in areas that remained formally unchanged.

Despite these limitations, this study provides valuable insights into child development trends and disparities. The findings contribute to evidence-based policymaking and the development of targeted intervention strategies.

## Conclusion

This study highlights that sociodemographic disparities in early childhood development are both dynamic and context-sensitive. Disparities associated with maternal characteristics, such as education, marital status, employment, maternal age, and country of birth, widened considerably during the reporting period, particularly in 2024. This widening coincided with two major national disruptions: the COVID-19 pandemic and the outbreak of war in late 2023. These overlapping crises appear to have amplified existing inequalities among subpopulations of mothers facing structural vulnerabilities. The most pronounced widening was observed in the language and social domains, underscoring the critical role of maternal background in buffering or intensifying developmental risks under conditions of systemic strain.

In contrast, disparities related to ethnicity and socioeconomic status remained relatively stable, and in some cases even narrowed, throughout the study period. In 2024, despite the significant disruptions, milestone failure rates among low-SES and minority ethnic populations did not deteriorate to the same extent observed in groups defined by maternal disadvantage. This may reflect the effect of targeted interventions, enhanced community resilience or improved service access among historically underserved populations.

These findings suggest that maternal-level risk factors may be particularly sensitive to contextual shocks. They highlight the need for equity-oriented developmental surveillance systems that remain robust during crises while being responsive to maternal profiles in order to guide more effective and inclusive intervention strategies.

## Declarations

### Ethics approval and consent to participate

All methods were performed in accordance with the ethical standards as laid down in the Declaration of Helsinki and its later amendments or comparable ethical standards. The study protocol was approved by the Soroka Medical Center institutional review board, approval number MHC-0006-18. An exemption of informed consent was granted by this ethics committee because the data were anonymous.

### Consent for publication

Not applicable.

### Availability of data and materials

The de-identified patient-level data used for this study contains sensitive information and therefore is not available outside the secured research environment of the Israel Ministry of Health. Summary aggregate level data and analysis code for this study can be made available upon reasonable request to the corresponding author.

### Authors’ contributions

IGi, GA, DRZ, RB, PA, MAT and YS conceptualized and designed the study. IGi, GA, IGo, DRZ, PA, MAT and YS contributed to data acquisition, analysis, or interpretation. IGi, GA and YS drafted the manuscript. IGi, GA, IGo, DRZ, RB, PA, MAT and YS revised the manuscript. IGi and IGo performed statistical analysis. DRZ, RB and MAT supplied technical or administrative support for the study. All authors read and approved the final manuscript.

## Data Availability

All data produced in the present study are available upon reasonable request to the authors

https://kinstitute.org.il/knowledge-center/

## List of abbreviations

CBS: Central Bureau of Statistics
FSU: Former Soviet Union
MCHC: Maternal child health clinic
OR: odds ratio
SES: Socioeconomic Status
THIS: Tipat Halav Israel Screening
VIF: variance inflation factor

## Acknowledgements

This study was conducted with the help of TIMNA – a national research platform established by the Israel government to enable big-data studies combining de-identified health data from multiple organizations.

---

**Supplementary Figure 1.**
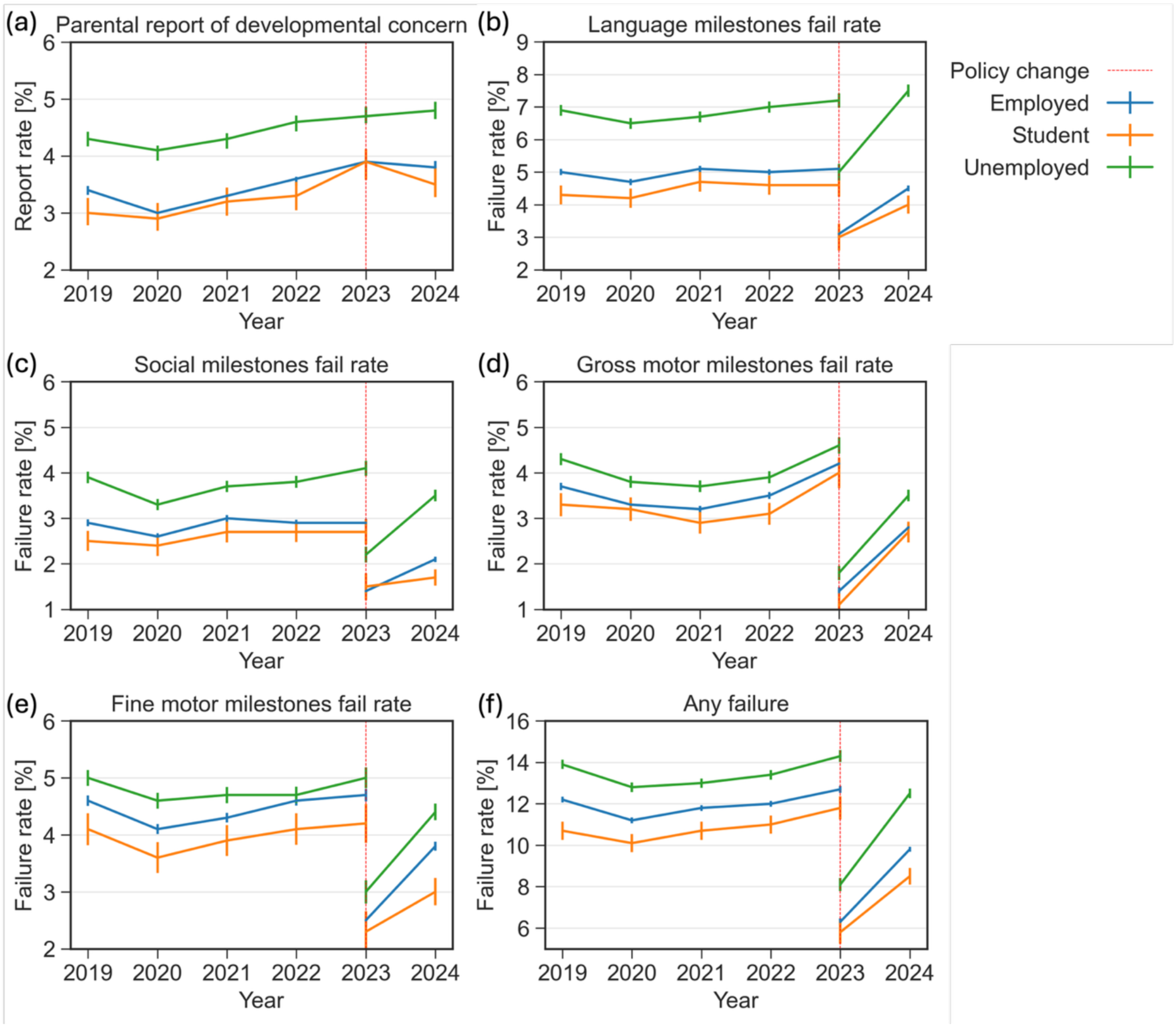
Reports of concern and failure rates in various developmental domains stratified by maternal employment status between 2019-2024. (a) Parental report of developmental concern (b) Failure rate in the language domain (c) Failure rate in the social domain (d) Failure rate in the gross motor domain (e) Failure rate in the fine motor domain (f) Failure rate in any developmental domain

**Supplementary Figure 2.**
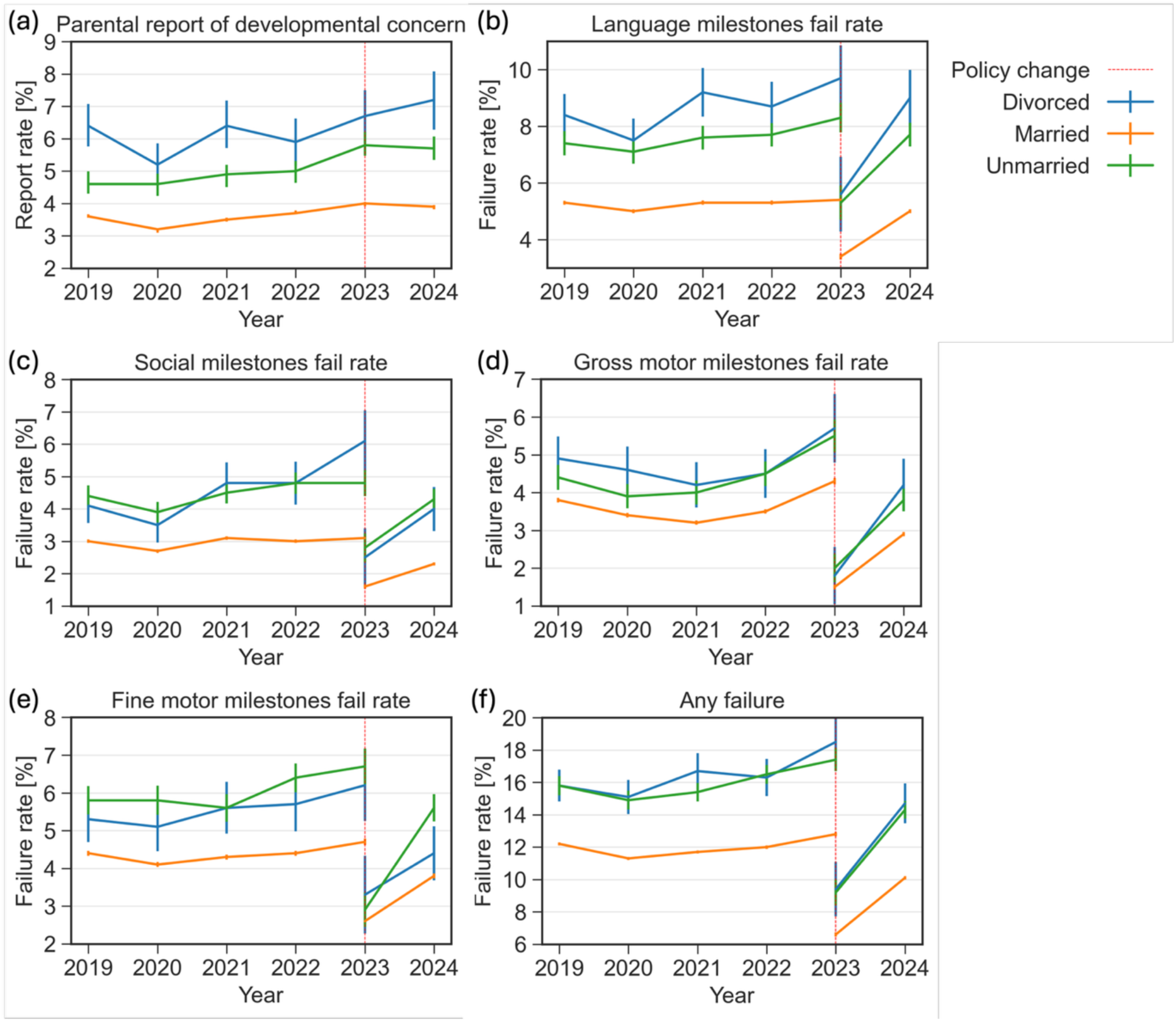
Reports of concern and failure rates in various developmental domains stratified by maternal marital status between 2019-2024. (a) Parental report of developmental concern (b) Failure rate in the language domain (c) Failure rate in the social domain (d) Failure rate in the gross motor domain (e) Failure rate in the fine motor domain (f) Failure rate in any developmental domain

**Supplementary Figure 3.**
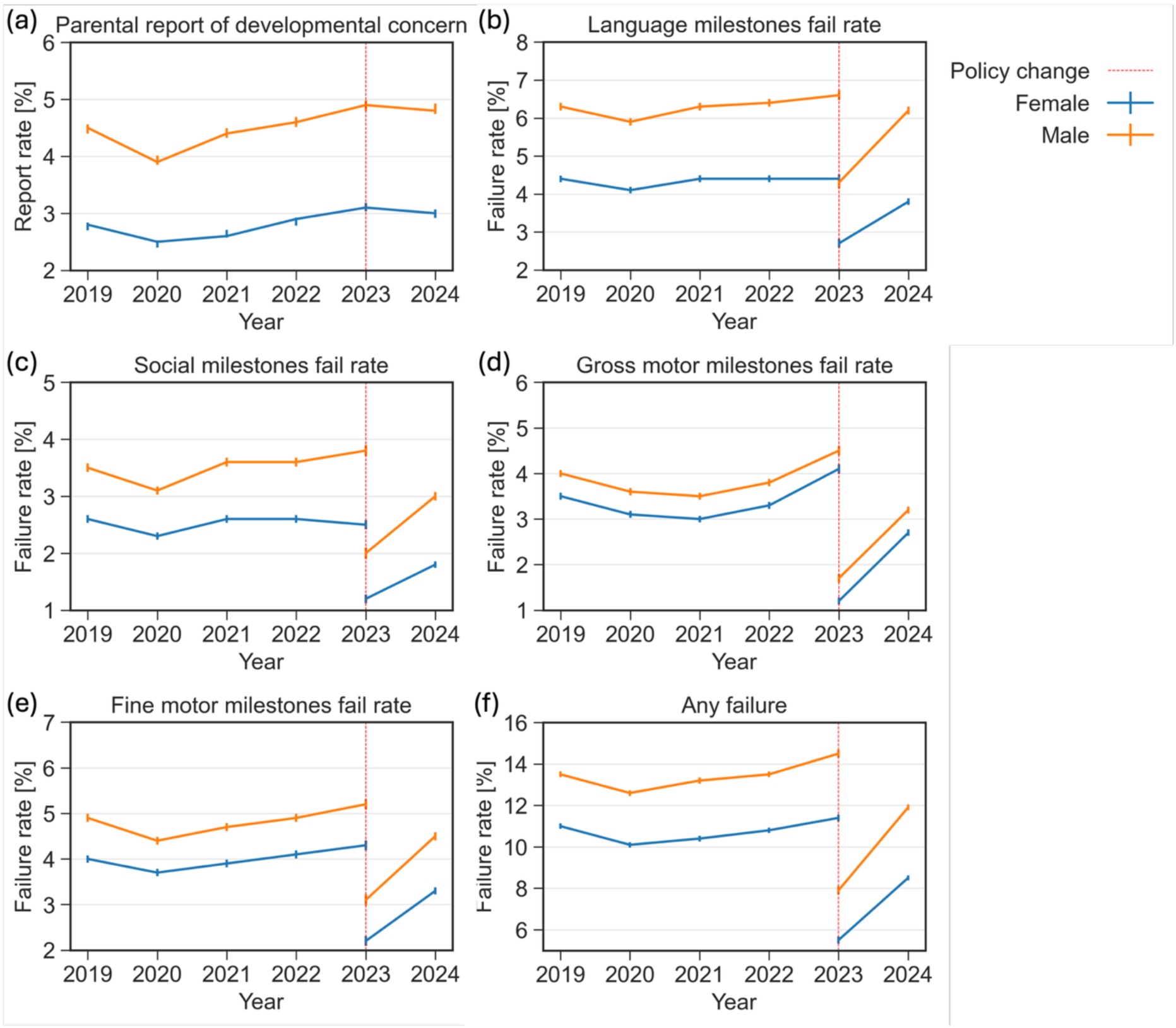
Reports of concern and failure rates in various developmental domains stratified by child’s sex between 2019-2024. (a) Parental report of developmental concern (b) Failure rate in the language domain (c) Failure rate in the social domain (d) Failure rate in the gross motor domain (e) Failure rate in the fine motor domain (f) Failure rate in any developmental domain

**Supplementary Figure 4.**
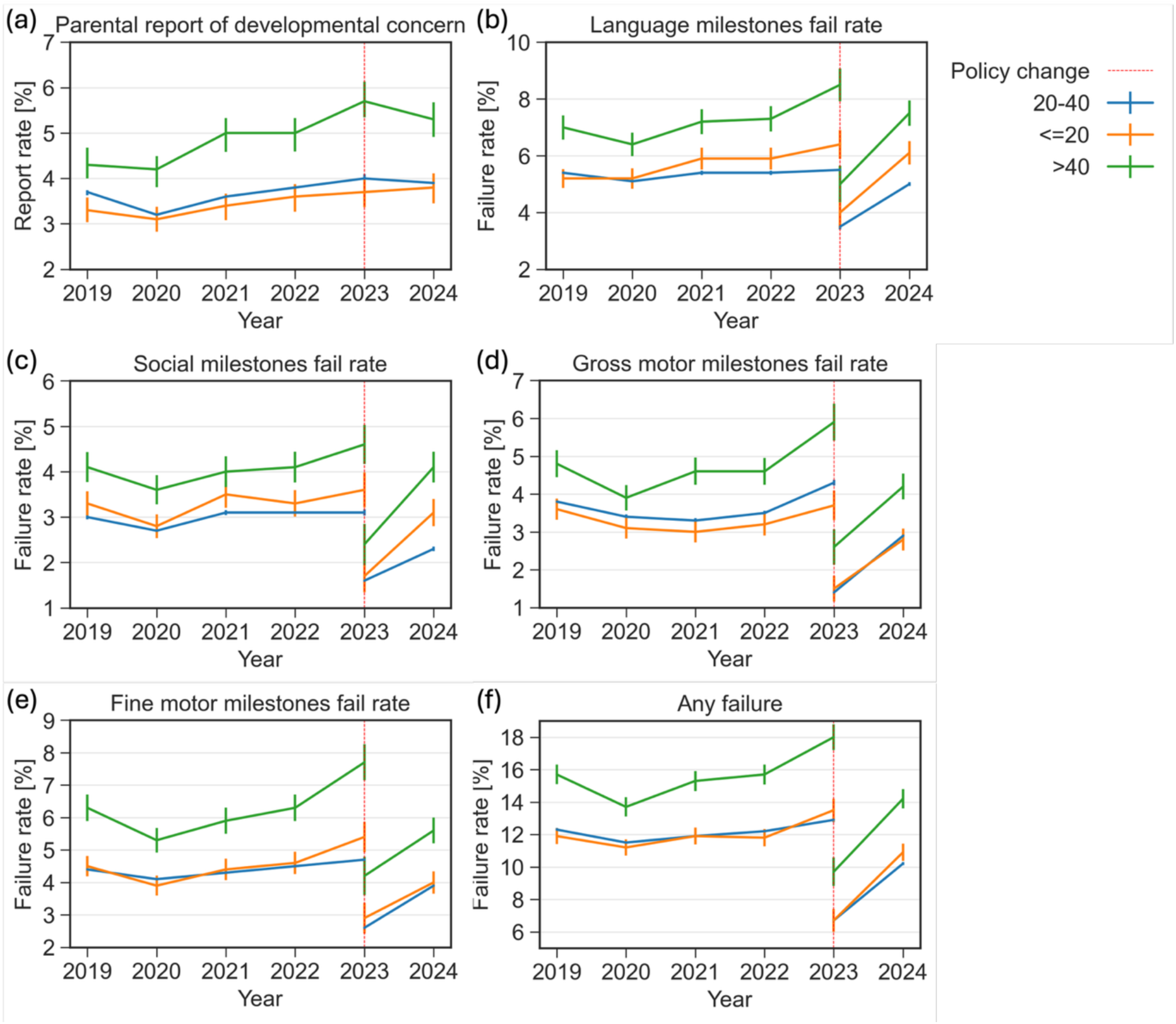
Reports of concern and failure rates in various developmental domains stratified by maternal age at childbirth between 2019-2024. (a) Parental report of developmental concern (b) Failure rate in the language domain (c) Failure rate in the social domain (d) Failure rate in the gross motor domain (e) Failure rate in the fine motor domain (f) Failure rate in any developmental domain

**Supplementary Figure 5.**
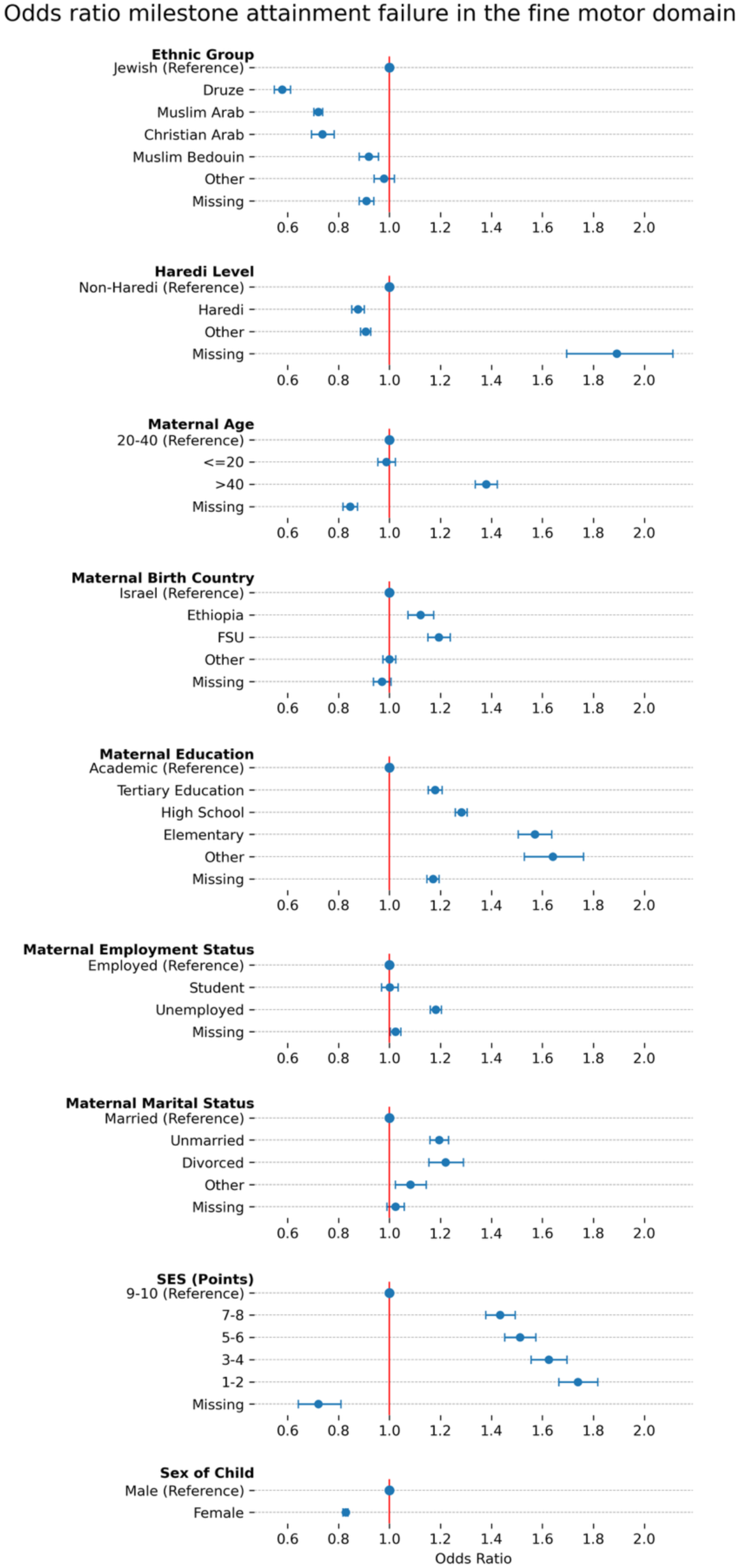
Multivariable analysis for failure in the fine motor domain for 2019-2022, where x axis is the odds ratio and y axis is the demographic variables.

**Supplementary Figure 6.**
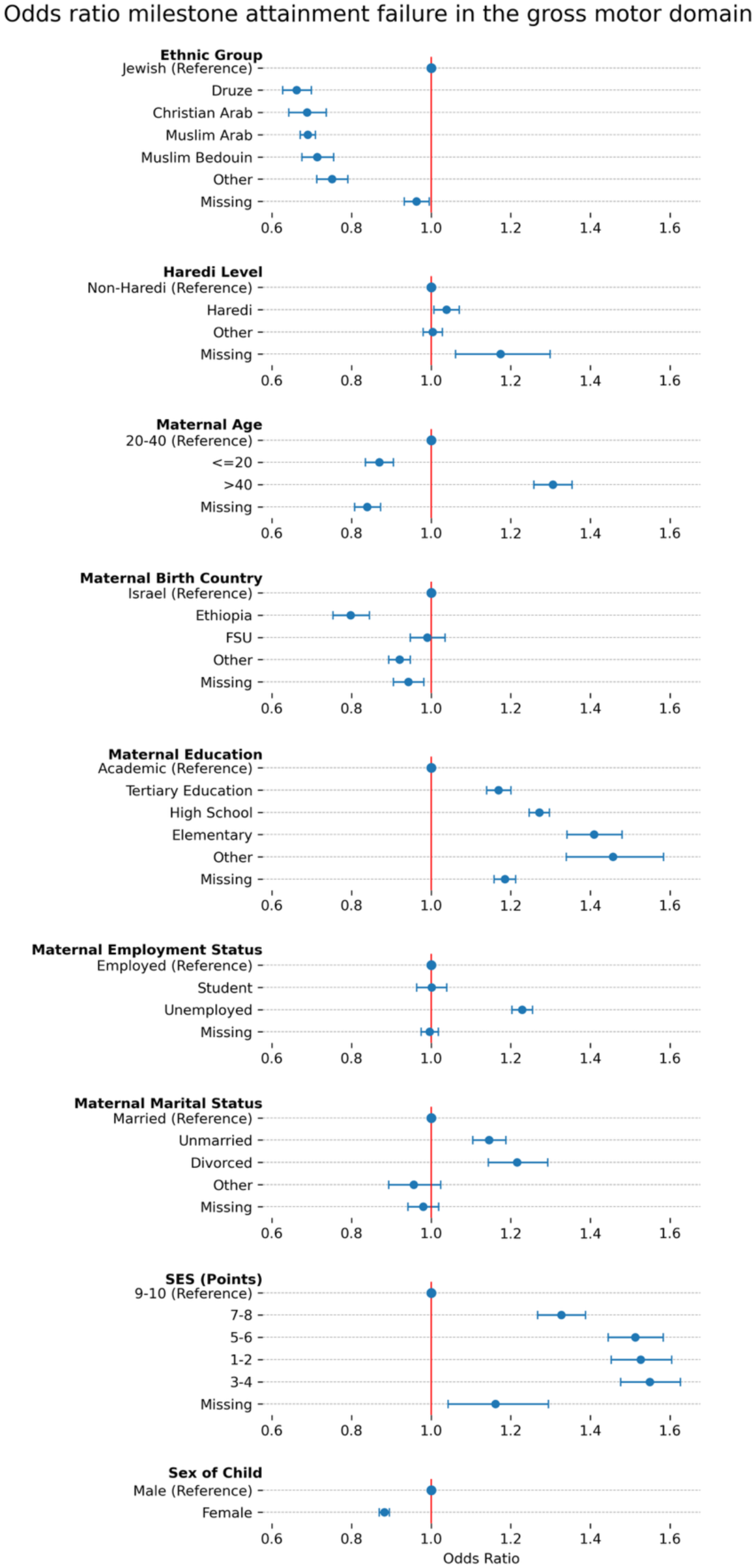
Multivariable analysis for failure in the gross motor domain for 2019-2022, where x axis is the odds ratio and y axis is the demographic variables.

**Supplementary Figure 7.**
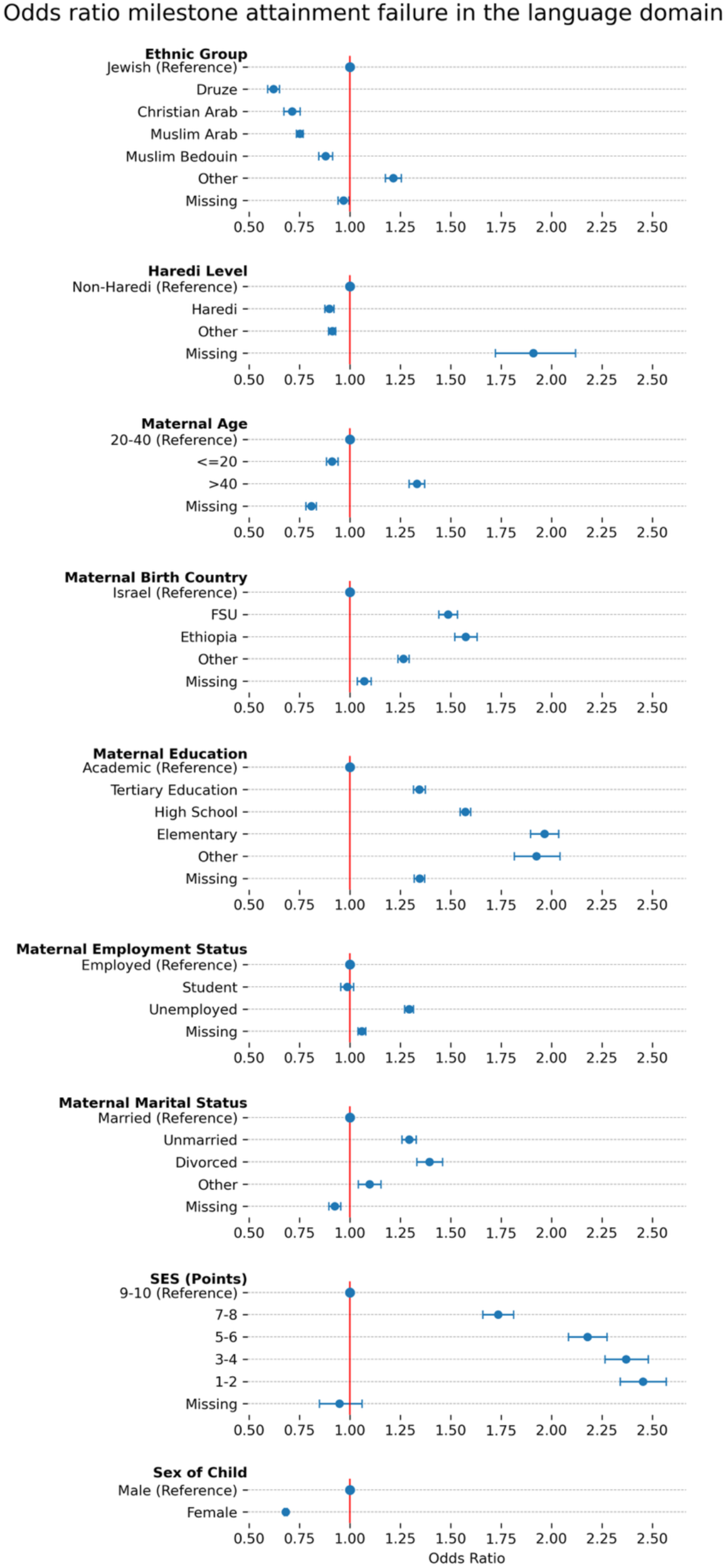
Multivariable analysis for failure in the language domain for 2019-2022, where x axis is the odds ratio and y axis is the demographic variables.

**Supplementary Figure 8.**
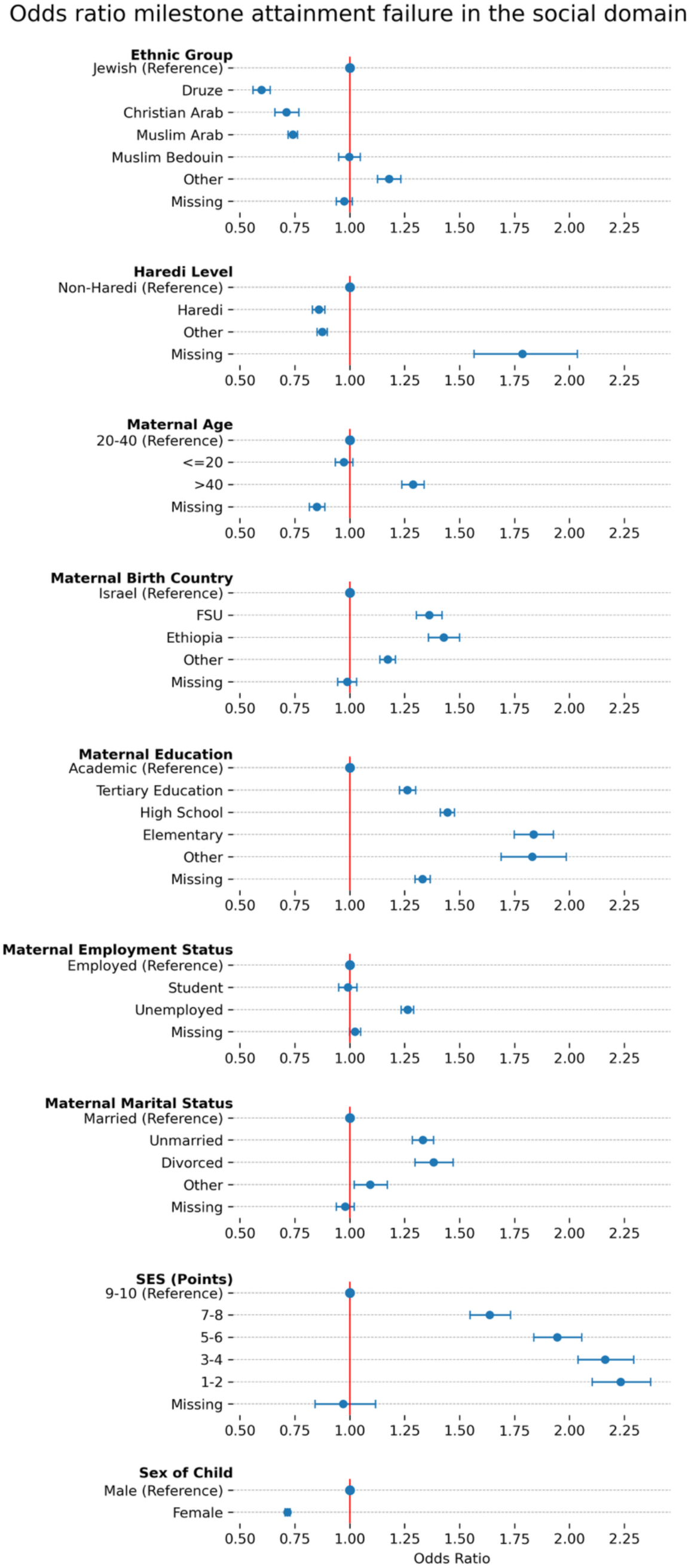
Multivariable analysis for failure in the social domain for 2019-2022, where x axis is the odds ratio and y axis is the demographic variables.

**Supplementary Figure 9.**
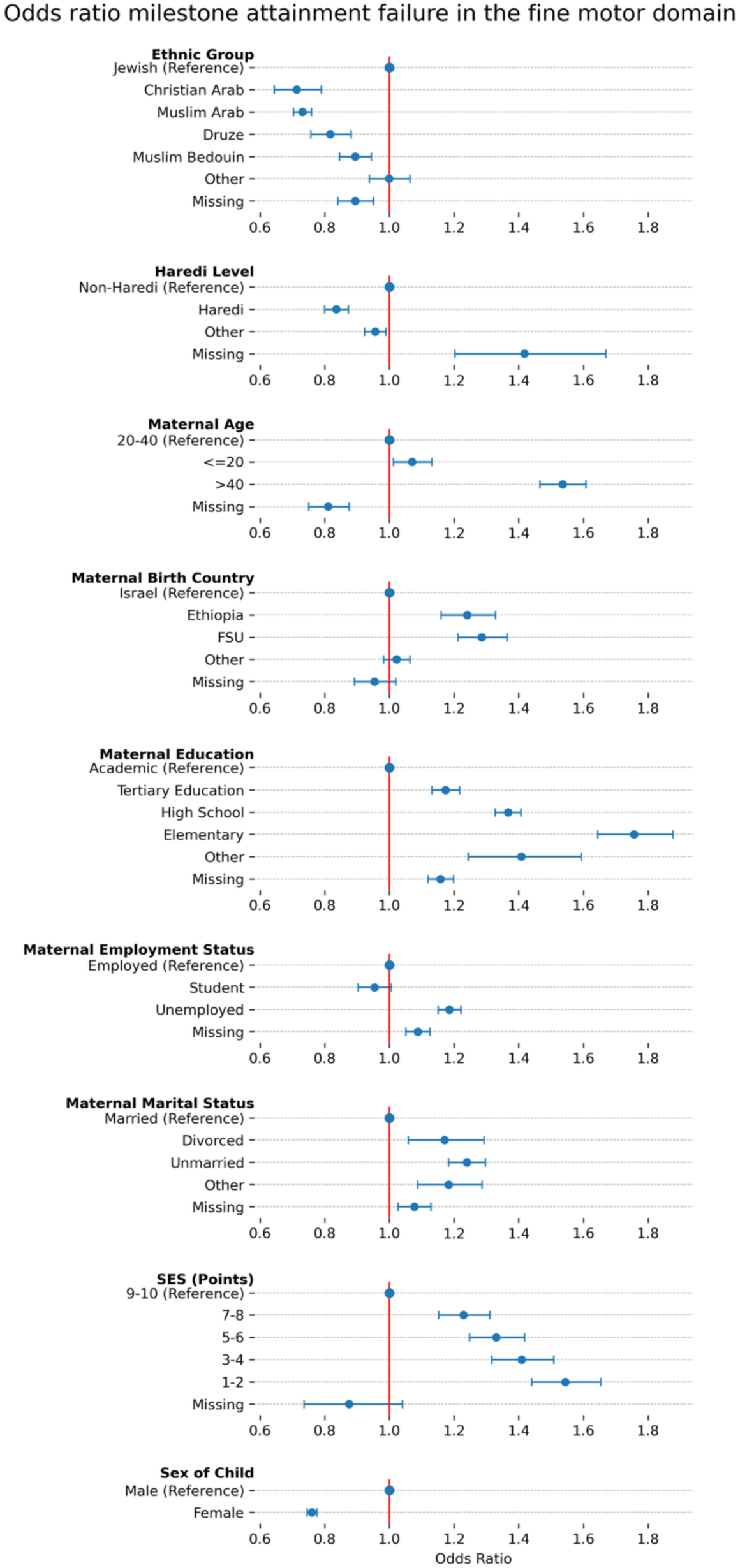
Multivariable analysis for failure in the fine motor domain for 2023-2024, where x axis is the odds ratio and y axis is the demographic variables.

**Supplementary Figure 10.**
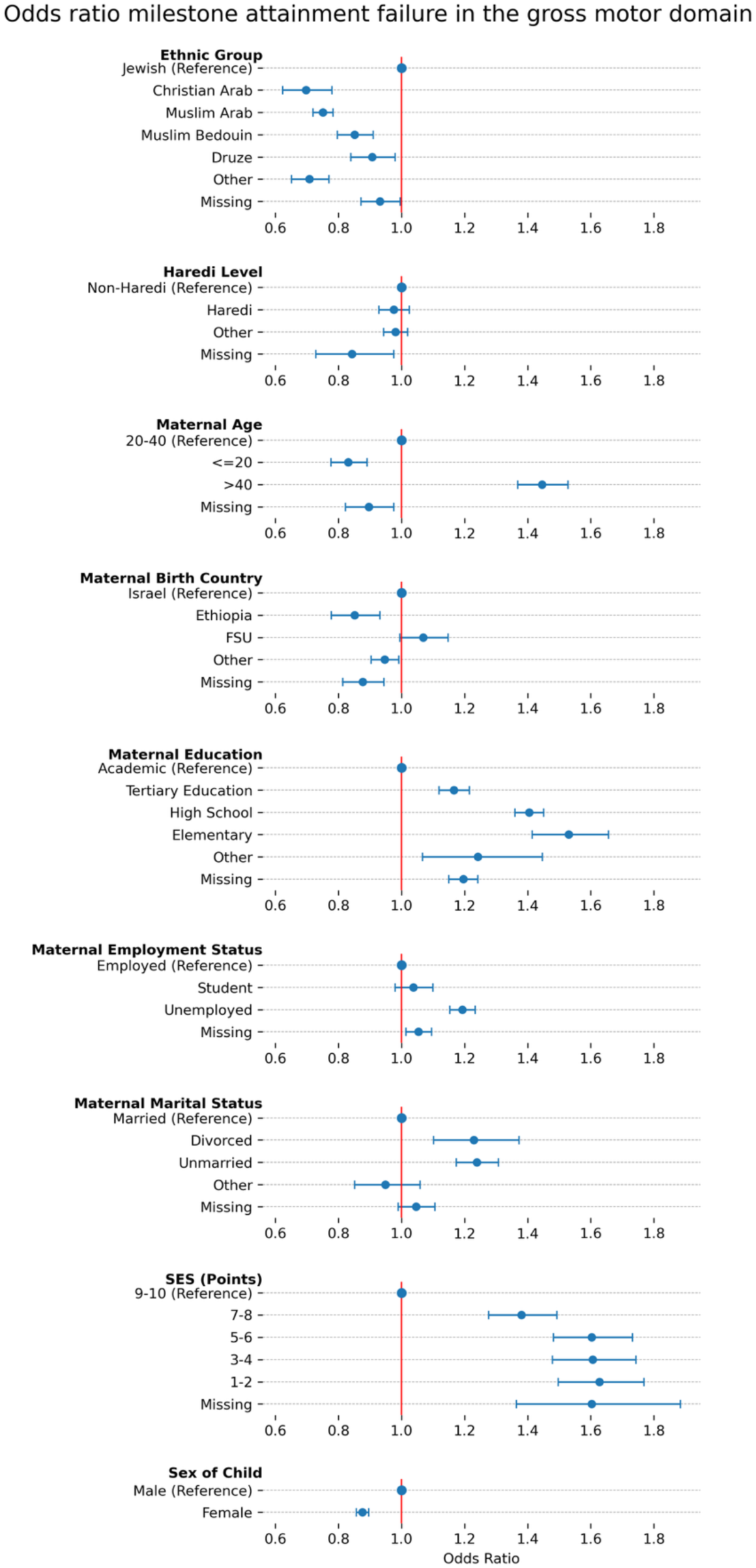
Multivariable analysis for failure in the gross motor domain for 2023-2024, where x axis is the odds ratio and y axis is the demographic variables.

**Supplementary Figure 11.**
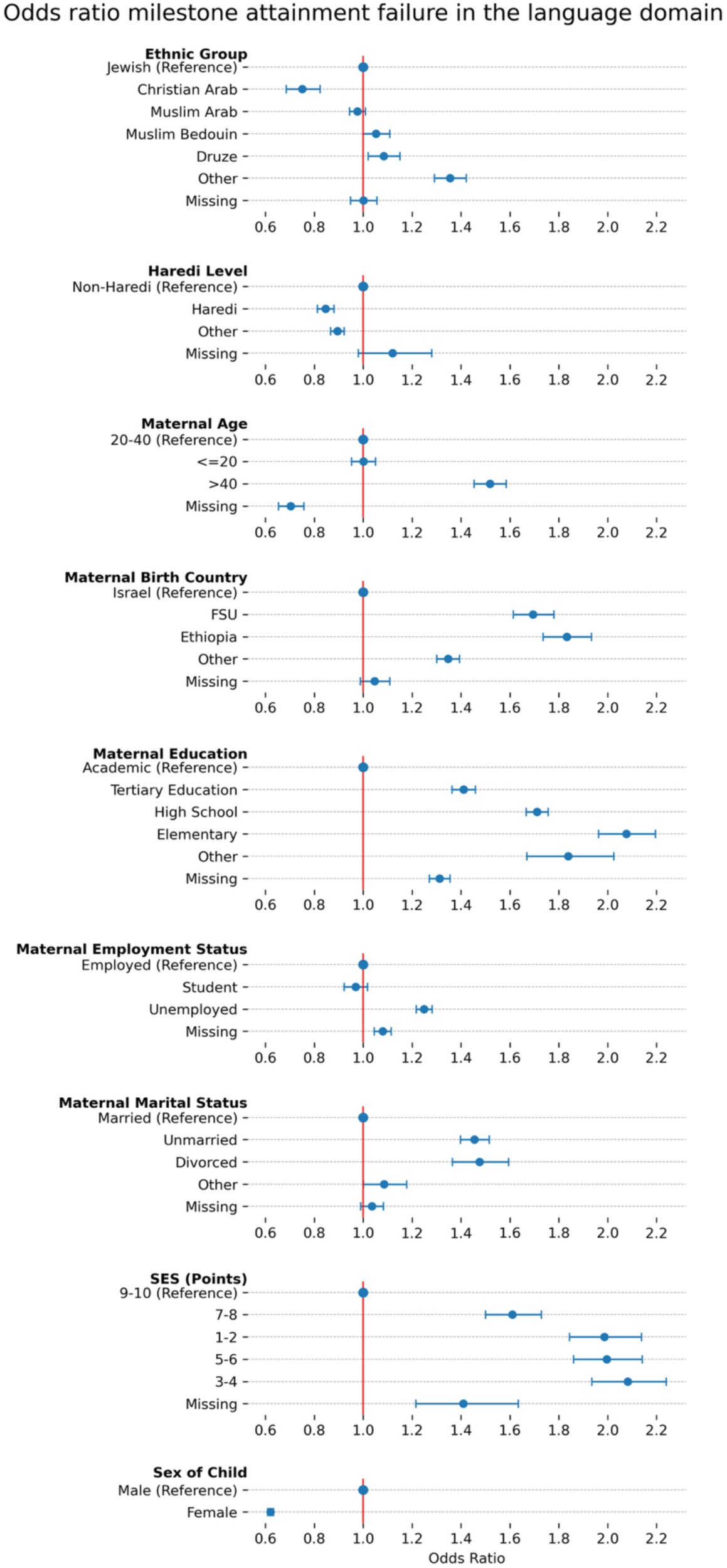
Multivariable analysis for failure in the language domain for 2023-2024, where x axis is the odds ratio and y axis is the demographic variables.

**Supplementary Figure 12.**
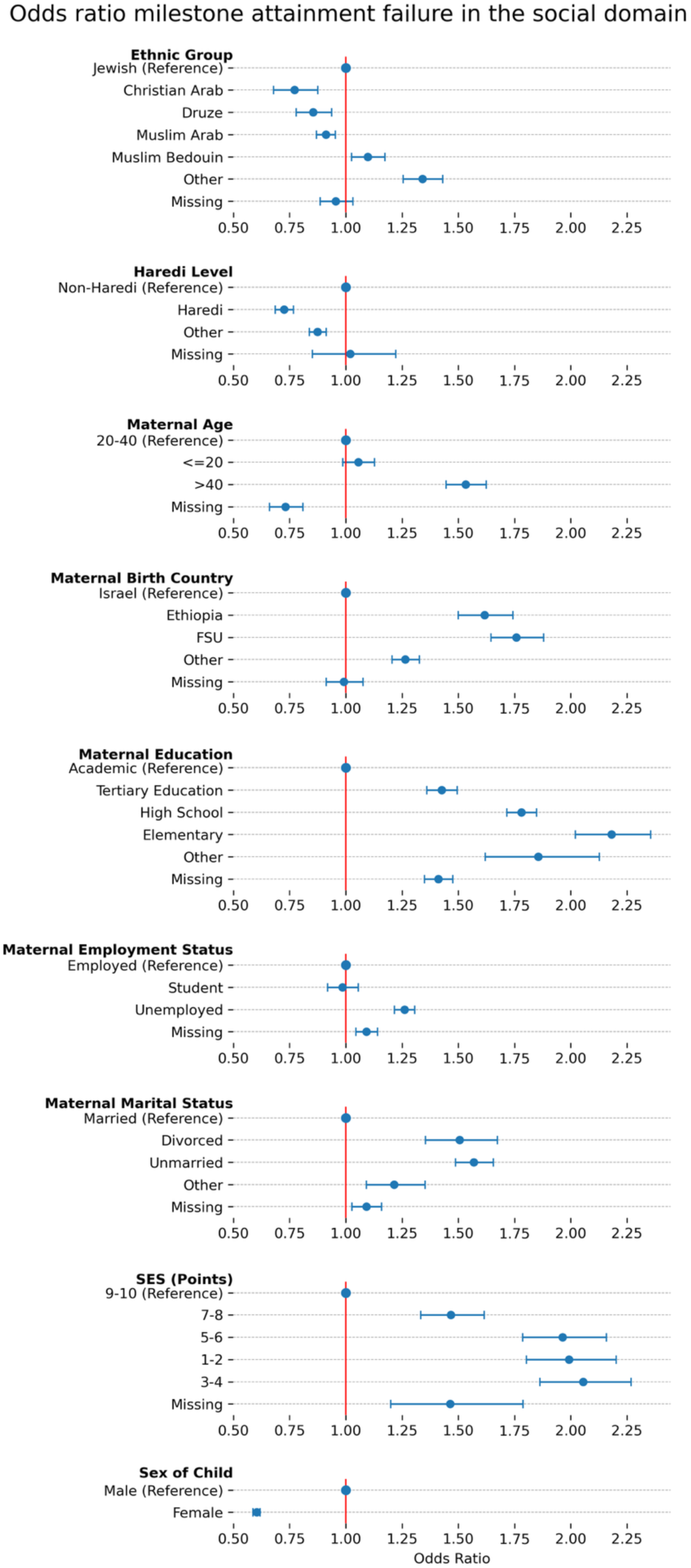
Multivariable analysis for failure in the social domain for 2023-2024, where x axis is the odds ratio and y axis is the demographic variables.

